# Genome-Wide Association Study of Non-Alcoholic Fatty Liver Disease Identifies Association with Apolipoprotein E

**DOI:** 10.1101/2021.05.05.21256592

**Authors:** Cameron J Fairfield, Thomas M Drake, Riinu Pius, Andrew D Bretherick, Archie Campbell, David W Clark, Jonathan A Fallowfield, Caroline Hayward, Neil C Henderson, Peter K Joshi, Nicholas L Mills, David J Porteous, Prakash Ramachandran, Robert K Semple, Catherine A Shaw, Cathie LM Sudlow, Paul RHJ Timmers, James F Wilson, Stephen J Wigmore, Ewen M Harrison, Athina Spiliopoulou

## Abstract

**Background & Aims:** Genome-wide association studies (GWAS) have identified several risk loci for non-alcoholic fatty liver disease (NAFLD). Previous studies have largely relied on small sample sizes and have assessed quantitative traits. We performed a case-control GWAS in the UK Biobank using recorded diagnosis of NAFLD based on diagnostic codes recommended in recent consensus guidelines.

**Approach & Results:** We performed a GWAS of 4,761 cases of NAFLD and 373,227 healthy controls without evidence of NAFLD. Sensitivity analyses were performed excluding other co-existing hepatic pathology, adjusting for BMI and adjusting for alcohol intake. 9,723,654 variants were assessed by logistic regression adjusted for age, sex, genetic principal components and genotyping batch. We performed a GWA meta-analysis using available summary association statistics from two previously published case-control GWAS of NAFLD. Six risk loci were identified (P<5*10^(−8)) of which one is novel in GWAS (rs429358 in APOE) and five are known (PNPLA3, TM6SF2, GCKR, MARC1 and TRIB1). Rs429358 (P=2.17*10^(−11)) is a missense variant within the APOE gene determining ⍰4 vs ⍰2/⍰3 alleles. All loci retained significance in sensitivity analyses without co-existent hepatic pathology and after adjustment for BMI. PNPLA3 and TM6SF2 remained significant after adjustment for alcohol (alcohol intake was known in only 158,388 individuals) with others demonstrating consistent direction and magnitude of effect. All 6 loci were significant on meta-analysis including APOE P=3.42*10^(−13) with consistent direction and magnitude of effect in all 6 loci in all three studies. The ⍰4 allele of APOE offered protection against NAFLD (odds ratio for heterozygotes 0.84 [95%CI 0.78-0.90] and homozygotes 0.64 [0.50-0.79]).

**Conclusions:** This GWAS demonstrates that the *∈*4 allele of *APOE* is strongly associated with protection against NAFLD.

---

Non-alcoholic fatty liver disease (NAFLD) is the commonest form of metabolic disease worldwide with an estimated prevalence of 24%, which appears to be increasing in all populations (1). NAFLD has an even higher prevalence in those with other forms of metabolic disease including obesity, type 2 diabetes mellitus and hyperlipidemia which are rising in prevalence meaning the prevalence of NAFLD is also anticipated to rise (1).

NAFLD covers a spectrum of disease severity including elevated hepatic triglyceride content (isolated steatosis), inflammation (non-alcoholic steatohepatitis), fibrosis and cirrhosis and is associated with elevated risk of further morbidity. It has risen to the second most common indication for liver transplantation in the United States and carries a significantly elevated risk of hepatocellular carcinoma, cardiovascular disease, diabetes mellitus and all-cause mortality (1). However, the pathogenesis of NAFLD is complex and disease progression is highly variable. Despite contributing to significant morbidity and mortality there are no licensed pharmacological therapies for NAFLD and several agents under investigation have thus far shown limited effectiveness (2).

Development of NAFLD is influenced by both environmental and genetic factors, its heritability estimated at 22-50% (3). Several genome-wide association studies (GWAS) have been published examining various phenotype definitions for NAFLD or NAFLD severity encompassing radiological evidence of hepatic steatosis, visceral fat content, lean NAFLD (NAFLD in a non-obese individual), and histological severity of fibrosis. These studies have highlighted several loci and novel candidate mechanisms underlying NAFLD pathogenesis (4-17).

Studies using a case-control GWAS methodology have identified five loci associated with NAFLD at genome-wide significance (P<5*10^−8^; MARC1, GCKR, HSD17B13, TM6SF2 and PNPLA3) (6-8,11-14,17). Known loci are shown in Supplementary Materials 6 (Supplementary Tables 1 and 2). Most case-control analyses have relied on histological and radiological confirmation of NAFLD limiting the sample size in these studies. Use of routinely-collected administrative data allows for larger cohorts and reduces the need for invasive or expensive investigations such as biopsy or magnetic resonance imaging (MRI) (18). One published GWAS relied on natural language processing to identify NAFLD cases from electronic health records (EHR) results and found significant associations at loci previously identified in cohorts with complete histological classification (12). Studies such as the UK Biobank (UKB) have extensive genotypic data linked to hospital and primary care discharge codes.

A subgroup of 14,440 participants in UKB has been analysed in a published GWAS focusing on MRI-based measures of steatohepatitis and fibrosis (15). Separately a case-control for all-cause cirrhosis using several sources including UKB has been published with an additional single variant analysis for NAFLD at rs2642438 using only two ICD10 diagnostic codes for NAFLD (K76.0 and K76.5) (14). There are no published case-control GWAS using administrative data based on recent consensus recommendations (18). We therefore undertook a GWAS of participants with recorded diagnostic codes attributed to NAFLD compared to healthy controls.

## Methods

### Study Populations

#### UK Biobank

The UKB is a prospective cohort study with 502,616 participants recruited between 2006 and 2010 from across the UK. Participants were men and women aged between 37 and 73 years at recruitment. Initial assessment involved self-completed touch-screen questionnaire, computer-assisted interview, physical and functional measures and collection of blood samples. Additional data has been generated from health record linkage to national registries and hospital discharges. Over 2.5 million hospital admissions were available for analysis. Additional primary care records were made available for 270,000 participants.

The recruitment process, consent process and data collection has been described extensively elsewhere (19-21). Data is available to researchers after a two-stage online application process. The UKB received ethical approval (research ethics committee reference 11/NW/0382). UKB data access was approved under projects 30439 (phenotype data) and 19655 (genotype data).

#### Generation Scotland: Scottish Family Health Study

Generation Scotland: Scottish Family Health Study (GS-SFHS) is a family-based genetic epidemiology study with DNA and socio-demographic and clinical data covering 24,096 volunteers across Scotland aged 18-98 years, from 2006 to 2011. Participants were identified within general practices in Scotland and were aged between 35 and 55 years with at least one first degree relative aged 18 years or over. Participants underwent baseline interview with a blood sample collected and stored for genotyping. Records for individuals were also linked to hospital- and community-based records using their community hospital index number provided to all citizens in Scotland registered with a general practice. A full description of the GS-SFHS protocol has been published previously (22,23). Ethical approval was granted by NHS Tayside Research Ethics Committee (REC reference number 05/S1401/89).

The study protocols conformed to the ethical guidelines of the 1975 Declaration of Helsinki.

### Identification of Non-Alcoholic Fatty Liver Disease

#### UK Biobank

NAFLD was defined as any hospital admission with an ICD-9 or ICD-10 code relating to NAFLD or any primary care encounter with a Read code relating to NAFLD. Controls were defined as those without NAFLD diagnoses. Whilst a diagnostic code should represent genuine NAFLD diagnoses, it is possible that the diagnostic code was incorrectly generated despite the presence of alternative hepatic pathology such as alcohol-related liver disease or viral hepatitis. A second cohort was therefore derived in which alternative hepatic pathology was excluded, remaining NAFLD cases were considered as cases, and all remaining healthy participants without any hepatic pathology served as controls. Diagnostic codes for identification of NAFLD and exclusion of differential diagnoses followed recent expert consensus guidelines (18). A full list of the relevant ICD and Read codes, in line with these recommendations, are available in Supplementary Materials 1.

#### Generation Scotland

NAFLD was defined as any hospital admission or primary care consultation resulting in generation of an ICD code or Read code relating to NAFLD. The codes and pathologies that were excluded were the same as in the UKB.

### Genotyping and Blood Sampling

For the UKB, collection of blood for genotyping was performed at study enrolment for approximately 470,000 individuals, the remainder were excluded due to insufficient sample volume. The cardiorespiratory-focused Affymetrix UK BiLEVE Axiom array was used in 50,000 participants and the Affymetrix UKB Axiom array was used for the remaining participants; the arrays were over 95% similar. Genotyping was performed in 106 batches (4000-5000 individuals per batch). Approximately 900,000 SNPs were directly genotyped and subsequent imputation resulted in 93 million SNPs for assessment. The full Affymetrix protocol and description of quality control performed prior to release of genotyped and imputed data to researchers have been published previously (19).

For GS-SFHS, collection of blood samples and genotyping was performed at study enrolment with 20,032 having DNA extracted via blood or saliva. Genotyping was performed on an Illumina HiScan platform and genotypes were called using GenomeStudio Analysis software v2011.1. Genotypes were imputed using the Haplotype Reference Consortium reference panel (HRC.r1-1) via the Sanger Imputation Server pipeline (https://imputation.sanger.ac.uk). A total of 24,161,581 SNPs were available for analysis. Genotype data covering each of the identified loci in the UKB GWAS were extracted from GS-SFHS.

### Genome-wide Association Study

#### Initial Analysis

Genotyped and imputed SNPs were analysed. Participants of self-reported European ancestry were considered eligible for inclusion. Outliers for heterozygosity and unexpected runs of homozygosity were excluded. One participant from each pair of related individuals in the UKB (Kinship > 0.0884) was excluded (24).

Association with NAFLD was analysed using logistic regression adjusted for age, sex, the first 20 genetic principal components and batch with batch included as a random effect. Imputation dosage was used for imputed SNPs.

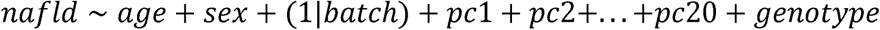

Quality control to exclude SNPs with low minor allele frequency (MAF<0.01) low imputation quality score (INFO<0.3) and deviation from Hardy-Weinberg Equilibrium (HWE: P<5*10^−6^) were applied. Genome-wide significance was determined as P<5*10^−8^ in the UKB and P<0.0083 in the GS-SFHS replication cohort (P value of 0.05 subjected to Bonferroni correction for each locus tested). Replication was undertaken by analysing the lead SNP within each locus for the GS-SFHS study or a linkage disequilibrium (LD) proxy. A power calculation was performed for novel findings to determine the required number of cases and the power available in the GS-SFHS cohort.

#### LD Clumping and Conditional Analyses

SNPs showing genome-wide significance (P<5*10^−8^) were considered significant. LD clumping was performed using the Functional Mapping and Annotation of Genome-Wide Association Studies (FUMA) web application v1.3.5d (https://fuma.ctglab.nl/) (25). Loci were established for lead SNPs with a minimum distance of 250Kb between loci and using an r^2^ < 0.25 to indicate independent SNPs within the same locus. Full details of the parameters passed to FUMA are available in Supplementary Materials 3A.

Each locus was re-analysed whilst conditioning on the lead SNP and further signals with genome-wide significance were identified. This process was repeated until no remaining SNPs reached genome-wide significance.

#### Population Stratification

The genomic inflation factor λ_gc_ was calculated using the summary association statistics. Evidence of test statistic inflation in λ_gc_ was investigated with linkage disequilibrium score regression (26).

### Sensitivity Analyses

Sensitivity analyses were conducted to ensure the robustness of the case definition. First, the analysis was conducted at each of the NAFLD-susceptibility loci after exclusion of individuals with alternative hepatic pathology as described above. Second, each of the analyses was conducted adjusting for body mass index (BMI) as a covariate. Third, the analysis was conducted adjusting for estimated consumption of alcohol (units per day). The alcohol estimate was derived from a 24-hour dietary recall questionnaire during online follow-up and was available for around 40% of participants. The logistic regression methodology for the sensitivity analyses was identical to the main GWAS.

An additional analysis in the UKB cohort was undertaken to assess the relationship between apolipoprotein E (*APOE*) genotype and NAFLD based on the GWAS results (one lead SNP was a missense mutation within *APOE*). Allelic status was determined using rs429358 and rs7412, which were both directly genotyped. Relationship was investigated by χ^2^ test and logistic regression. See Supplementary Materials 5 for methods explaining determination of *APOE* alleles.

### Association of NAFLD-susceptibility Loci with Phenotypic Traits

NAFLD is strongly associated with obesity, hyperlipidemia, hyperglycemia, inflammation and deranged liver function tests, in particular alanine aminotransferase (ALT) (27,28). Associations with serum biochemistry values for each identified variant were assessed in an age- and sex-adjusted linear regression model. Serum ALT, aspartate aminotransferase (AST), gamma-glutamyl transferase (GGT), alkaline phosphatase (ALP), total cholesterol (TC), low-density lipoprotein cholesterol (LDL-C), high-density lipoprotein cholesterol (HDL-C), triglycerides, apolipoprotein A and B (ApoA and ApoB), lipoprotein A (LipoA), glucose, glycated hemoglobin (HbA1c), C-reactive protein (CRP), BMI, waist circumference, hip circumference and waist-to-hip ratio were assessed. Each trait was assessed visually via histogram and log-transformed in the case of skewed distribution.

A linear regression adjusted for age, sex, the first 20 genetic principal components and genotyping batch was undertaken for the association of NAFLD-susceptibility variants with the FIB-4 score and the NAFLD fibrosis score, both validated non-invasive measures associated with NAFLD-related fibrosis (29,30).

### Genome-wide Association Meta-analysis

Two NAFLD case-control GWAS with available summary association statistics (Namjou et al. (12): 1,106 cases and 8,571 controls; and Anstee et al. (13): 1,483 cases and 17,781 controls) from independent populations of European ancestry were also assessed. Namjou et al. report on participants from American health centers and relied on natural language processing to ascertain cases of NAFLD from the EHR and, as such, is more closely related to the methodology used in this study. Namjou et al. performed a case-control logistic regression adjusted for age, sex, BMI, the medical center and the first three genetic principal components. Anstee et al. recruited participants from European tertiary liver centers with biopsy-proven NAFLD and compared them to population controls with a linear mixed model adjusting for sex and the first five genetic principal components before repeating the analysis as a logistic regression. Identified lead SNPs from the UKB GWAS were inspected in these studies for direction and magnitude of effect.

Meta-analysis was conducted using METAL (31) with an inverse-variance fixed effects meta-analysis. Whilst between-study heterogeneity was expected, the fixed effects model was used in preference to the random effects model due to the low number of studies and anticipated deviation from the Gaussian distribution required for a random effects model (32).

### Plotting and Statistical Analysis of Results

Genomic analyses were performed using SNPtest (version 2.5.2), QCTOOLS (version 2.0.6) (33) and METAL (2018 version) on the University of Edinburgh Linux high-performance compute cluster. Post-GWAS analysis, regression analyses and plotting were performed using R version 3.6.3 (34). Methods and results are reported in accordance with the STREGA guidelines (35) (see Supplementary Materials 8).

## Results

### NAFLD Cohorts

502,616 participants entered the UKB study with 377,998 taken forward for GWAS. A history of NAFLD was present in 4,761 with 373,227 controls. After exclusion of alternative hepatic pathology there were 3,954 NAFLD cases and 355,942 controls. The median follow-up was 9.0 years (range 7.3-11.9). Compared to controls, cases were more likely to be male (51.0% vs 46.3%), older (mean 57.4 vs 56.9 years), heavier (mean 89.2 vs 78.2kg) and diabetic (32.9% vs 7.7%). The baseline characteristics are shown in Supplementary Materials 6 (Supplementary Table 3).

24,096 participants entered GS-SFHS with 6,317 taken forward for GWAS. A history of NAFLD was present in 67 with 6,250 controls (both without alternative pathologies). The mean follow-up was 11.2 years (range 9.6-14.7 years) (Figure 1).

**Figure 1:**
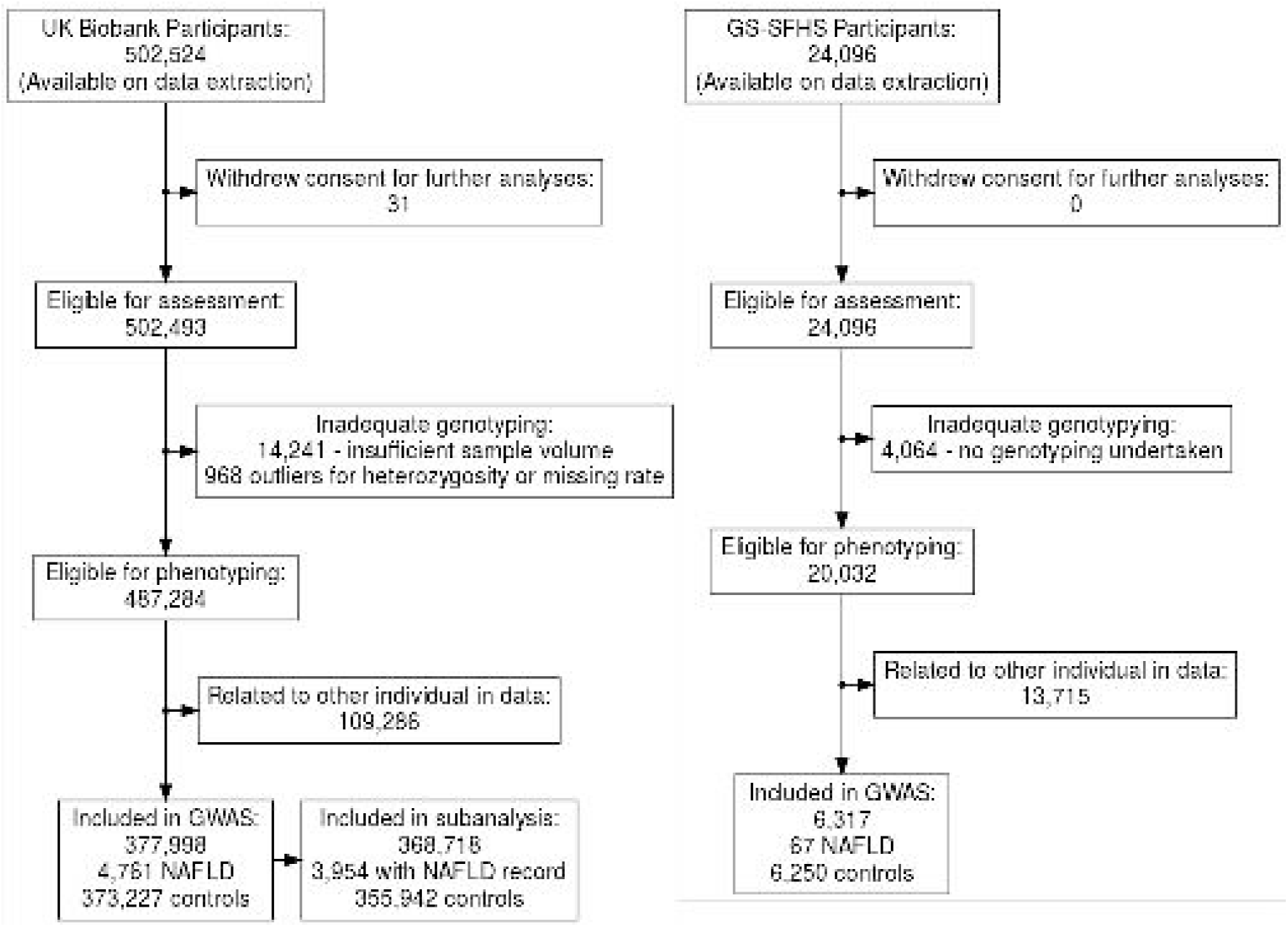
Flowchart of Participant Recruitment. Flowchart describing participant recruitment in UK Biobank and Generation Scotland: Scottish Family Health Study

### Genome-wide Association Study

#### Initial Analysis

460 SNPs were identified with genome-wide significant P values after removal of low-quality SNPs in the UKB (see Supplementary Materials 2). 1,313 SNPs demonstrated borderline significance (P<5*10^−5^). LD-clumping revealed 6 loci with at least one significant NAFLD-associated signal (see Supplementary Materials 3B). Significant associations were seen with rs2642442 (mitochondrial amidoxime reducing component 1 [*MARC1*] intron), rs1260326 (glucokinase regulator [*GCKR*] exon), rs17321515 (tribbles pseudokinase 1 [*TRIB1*] intergenic), rs73001065 (maternal-effect uncoordinated sister chromatid cohesion factor [*MAU2*] intron), rs429358 (apolipoprotein E [*APOE*] exon) and rs3747207 (patatin-like phospholipase domain-containing 3 [*PNPLA3*] intron). Rs73001065 is in strong linkage with the previously identified missense variant (rs58542926) within *TM6SF2* (R^2^=0.82) (11,13) which is thought to be the causal variant (36).

Of the 6 loci, 5 are known from previous case-control GWAS of NAFLD or related traits such as hepatic steatosis. One is a newly identified variant (rs429358-*APOE*) (Table 1). One locus (rs1260326) was replicated within the GS-SFHS cohort, 3 were replicated by Namjou et al. and all 6 by Anstee et al. (P < 0.0083; Table 2). GWAS meta-analysis resulted in broadly similar results with no additional loci reaching genome-wide significance. Direction and magnitude of effect were similar for all 6 loci other than rs73001065-C for which GS-SFHS demonstrated a non-significant reduction in NAFLD risk but with substantially wider confidence intervals. A power calculation (http://csg.sph.umich.edu/abecasis/cats/gas_power_calculator/) was conducted for the *APOE* locus using a desired P value of 0.0083, a disease prevalence of 1%, disease allele frequency of 84.8% and a relative risk of 1.21. This demonstrated a required sample with 2,500 cases to achieve a power of 80%.

Forest plots showing effect size in the four studies (UKB GWAS cohort, GS-SFHS replication cohort, Namjou et al. summary association statistics and Anstee et al. summary association statistics) along with a Manhattan plot from the GWAS meta-analysis are shown in Supplementary Materials 6 (Supplementary Figures 7 and 8).

One previously identified locus (rs13118664 (13)) did not reach genome-wide significance but did reach the replication threshold (P =0.004) with the G allele also demonstrating reduction in NAFLD risk in our study.

A Manhattan plot for association of variants with NAFLD in the UKB is shown in Figure 2.

**Figure 2:**
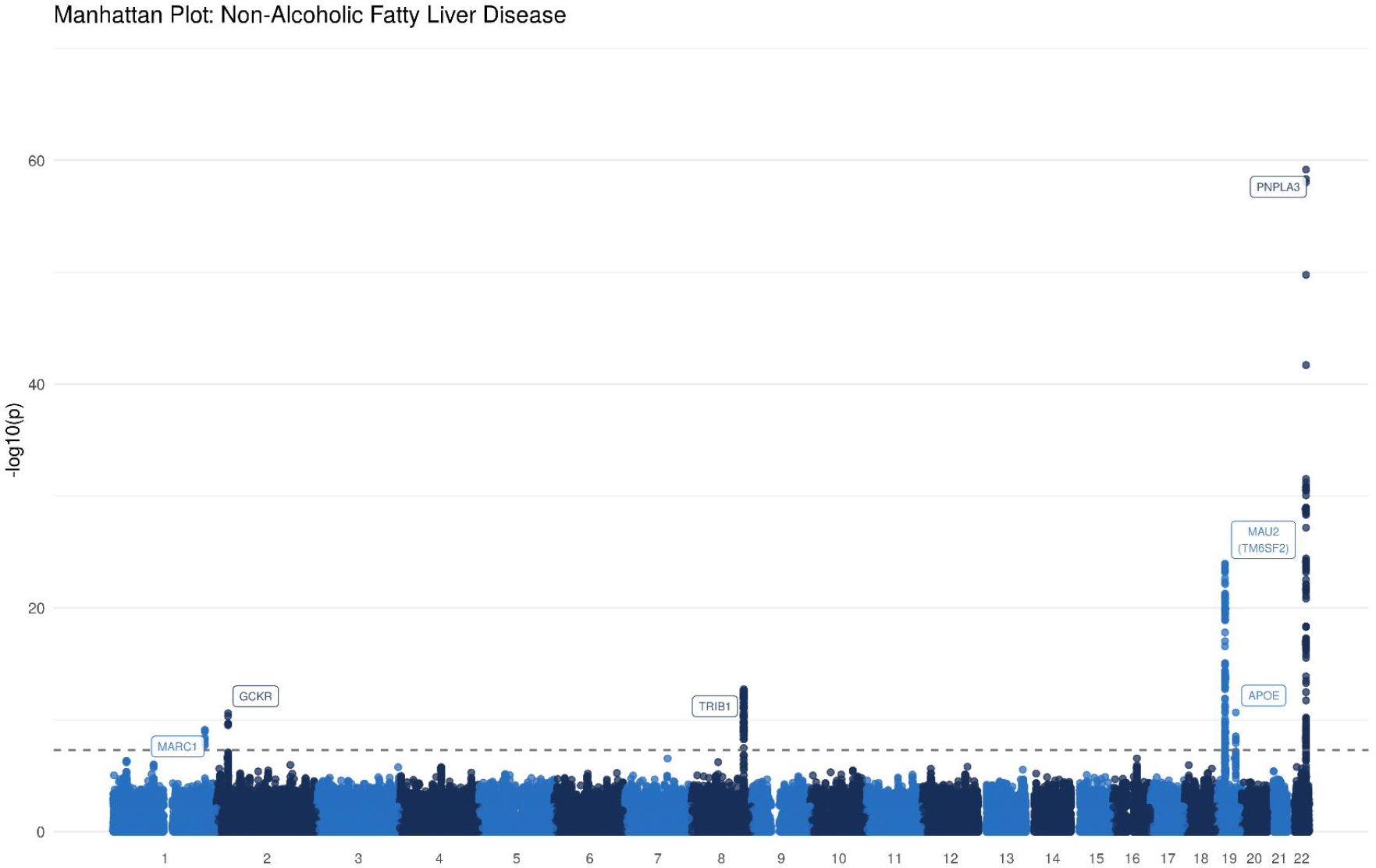
Manhattan Plot: Non-Alcoholic Fatty Liver Disease. Manhattan plot for the association with non-alcoholic fatty liver disease (4,761 cases and 373,227 controls). Each variant is plotted based on chromosome and position on the X-axis and -log10 P values on the Y-axis. The horizontal dotted line represents genome-wide significance (P = 5*10^−8^).

#### Conditional Analyses

After conditional analyses one locus was found to have a further independent signal. Rs182611493 within the *MAU2*/*TM6SF2* locus showed significant association (P=9.30*10^−13^) after conditioning on rs73001065 (see Supplementary Materials 4). The remaining 5 loci did not have any SNPs reaching genome-wide significance (P<5*10^−8^) after conditioning on the lead SNP.

### Population Stratification

After adjusting for the first 20 genetic principal components, there was evidence of test statistic inflation (λ_gc_=1.057). Inflation may be due to polygenicity rather than unmeasured population substructure (37). The linkage disequilibrium score regression intercept was 1.006 and the proportion of test statistic inflation ascribed to causes other than polygenicity was estimated to be 7.52% confirming that polygenicity is the main driver of test statistic inflation. The quantile-quantile plot is shown in Supplementary Materials 6 (Supplementary Figure 9).

### Sensitivity Analyses

Across all sensitivity analyses, the estimated genetic effects at each lead SNP had the same direction and broadly similar magnitude. All SNPs demonstrated a significant effect when alternative hepatic pathologies were excluded and when adjusting for BMI. Two SNPs retained significance after additionally adjusting for alcohol intake (rs73001065, rs3747207), with one retaining suggestive significance (rs17321515, P=1.39*10^−7^). The other three SNPs no longer retained suggestive significance although this is likely to be due to the greatly reduced sample size in those individuals who had completed the alcohol intake questionnaire (158,388 versus 377,998) with all 6 SNPs showing greatly attenuated significance. Visual inspection of the lattice plots showed that odds ratio estimates at each SNP were broadly similar with wider confidence intervals in the alcohol-adjusted analysis (see Supplementary Materials 6 – Supplementary Figure 6).

*APOE* genotype was significantly associated with NAFLD (χ^2^=2.49*10^−8^). The *∈*4 allele was strongly associated with reduced risk. The odds ratio for NAFLD risk for *∈*3/*∈*4 heterozygotes was 0.84 (95% CI 0.78-0.90) and for *∈*4/*∈*4 homozygotes 0.64 (0.50-0.79). The *∈*2 allele was not associated with any significant change in NAFLD risk. The odds ratio for NAFLD risk for *∈*2/*∈*3 heterozygotes was 1.02 (95% CI 0.93-1.11) and for *∈*2/*∈*2 homozygotes 0.76 (0.50-1.12). All odds ratios relate to the *∈*3/*∈*3 homozygote reference group. The results of the logistic regression are shown in Figure 3. See Supplementary Materials 5 for full results.

**Figure 3:**
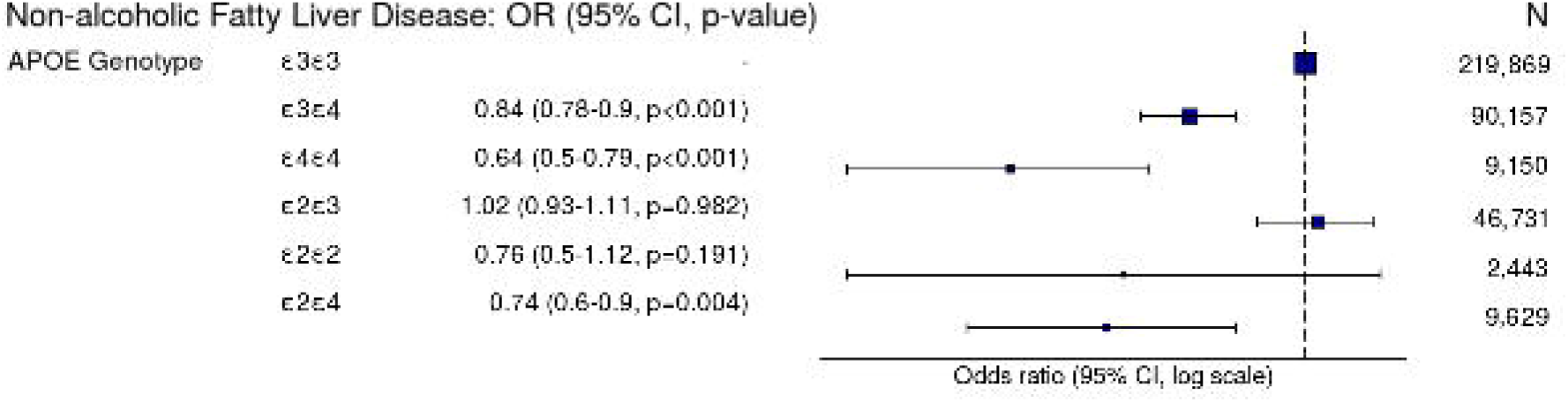
Odds Ratio Plot: Association of Non-Alcoholic Fatty Liver Disease and Apolipoprotein E Genotype. Odds ratio plot demonstrating odds of non-alcoholic fatty liver disease (NAFLD) by Apolipoprotein E (*APOE*) genotype. Each APOE genotype is compared to the ε3 homozygotes reference group. (Model adjusted for age, sex, genotyping batch and the first 20 genetic principal components).

### Association of NAFLD-susceptibility Loci with Phenotypic Traits

Assessment of serum lipids was undertaken for each lead SNP in all 377,998 individuals taken forward for GWAS. The NAFLD-susceptibility alleles were heterogeneous in their influence on serum lipids with most demonstrating reduced levels of TC and LDL other than rs17321515 (see Figure 4). Four variants including the novel variant were associated with elevated ALT (rs17321515, rs73001065, rs429358, rs3747207). Figures for the other serum biochemistry markers are available in Supplementary Materials 6 (Supplementary Figures 1-5).

**Figure 4:**
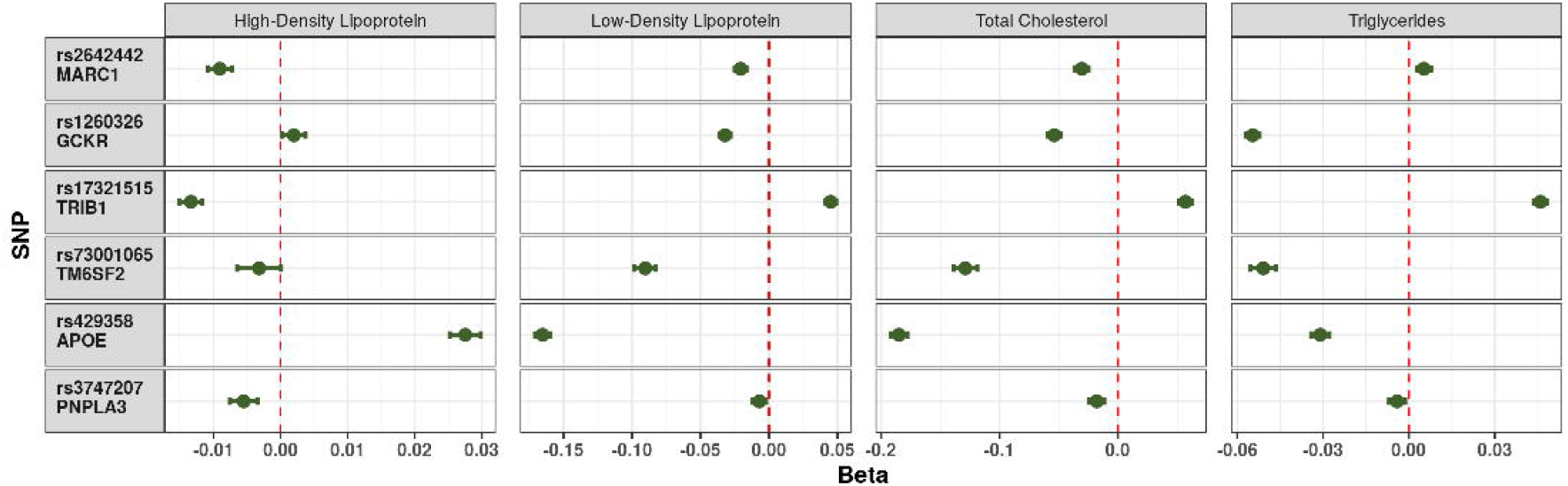
Forest Plots: Impact of Non-Alcoholic Fatty Liver Disease Susceptibility Loci on Serum Cholesterol. Impact of each NAFLD-susceptibility allele on the measured serum cholesterol fractions. Each point represents the beta-coefficient from an age- and sex-adjusted linear regression and the error bar represents the 95% confidence interval. Triglycerides were log-transformed prior to analysis. Chol - Total cholesterol; HDL - high-density lipoprotein; LDL - low-density lipoprotein; trigs – triglycerides.

The NAFLD-susceptibility variant within *APOE* was associated with decreased TC, LDL, triglycerides, ApoA, LipoA, HbA1c, ALT, ALP and GGT. The NAFLD-susceptibility variant was the major allele of rs429358-T (*∈*3).

In patients with NAFLD, only the locus with the strongest signal was associated with a change in FIB-4 Score on linear regression (rs3747207: Beta 0.08 [95% CI 0.03-0.14], p=0.001). The NAFLD fibrosis score was not significantly influenced by any of the NAFLD-susceptibility loci.

## Discussion

We performed a GWAS of NAFLD using 4,761 cases and 373,227 controls from the UKB study. We identified rs429358-C within *APOE*, a protein-altering variant, which is protective against NAFLD. We replicated all five previously identified loci in NAFLD case-control GWAS, of which four reached genome-wide significance and replicated a further locus associated with hepatic steatosis. We replicated our novel finding using summary association statistics published from independent populations.

Rs429358 is a missense variant within *APOE* which in combination with rs7412 defines the three main alleles of *APOE*, namely *∈*3, *∈*4 and *∈*2 (38). *APOE* plays various roles in peripheral lipid and lipoprotein metabolism and the three common alleles influence metabolic and cardiovascular disease and Alzheimer’s disease (39). The role of *APOE* in NAFLD has been examined previously in candidate-gene studies although has not previously been detected by GWAS. Whilst this manuscript was under preparation, an additional exome-wide array meta-analysis has been published in which rs2075650 in *TOMM40* (translocase of outer mitochondrial membrane 40) was identified. The authors suggest based on conditional analysis that rs429358 in *APOE* is the causal variant with the C allele conferring protection (40). Candidate genes studies have previously shown a decreased risk with the *∈*4 allele (41-43) and the *∈*2 allele (42,44) although some studies reported no difference in the risk of NAFLD (45,46). Elevated serum levels of *APOE* appear to correlate with higher fatty liver index regardless of genotype (46). Perhaps surprisingly, the *∈*4 allele may be associated with greater NAFLD fibrosis severity although this finding was made in a very small sample (47). The existence of additional populations demonstrating a similar association confirms that the *APOE* finding is likely to be a genuine association. The apparent lack of effect in some of the earlier studies is likely related to smaller sample sizes.

The mechanisms by which *APOE* influences NAFLD development remain unclear. There is a linear increase in both cardiovascular risk and serum LDL and TC with transition from *∈*2 to *∈*3 and from *∈*3 to *∈*4 (39). The association between NAFLD and cardiovascular and metabolic disease (1) is unlikely to be explained by *APOE* activity given the *∈*4 allele simulataneously offers protection against NAFLD and increases the risk of cardiovascular and metabolic disease. *APOE* influences hepatic very-low-density lipoprotein (VLDL) secretion (48). Apoe-deficient mice demonstrate reduced VLDL secretion and greater steatohepatitis severity. This is not corrected by APOE-producing bone marrow transplants with hepatic VLDL secretion remaining low, confirming that *APOE* plays an important role in liver autonomous VLDL secretion (49). The *∈*4 allele is associated with enhanced hepatic VLDL secretion and this may explain the association with hypertriglyceridemia and cardiovascular disease as well as protection against hepatic steatosis (50).

The other loci have been identified by previous GWAS demonstrating that the EHR-based approach generates results similar to histopathological and radiological methodologies.

The strengths of this study include using a larger sample size than previous studies, with greater power to detect association. The study also used a second cohort for external replication as well as available GWAS summary association statistics from other populations resulting in replication of all identified loci. The use of administrative records for identification of cases also demonstrates that this technique can be used to study NAFLD without the requirement for invasive procedures or radiological assessment. Our study has confirmed two loci identified using radiological assessment of NAFLD in a smaller subset of the UKB data (14,400 participants; *TM6SF2* and *PNPLA3*) (15) and three other SNPs identified in other GWAS. Furthermore, the strongest known signal at *PNPLA3* was verified in this study with a highly significant association. The study also benefits from demonstrating the robustness of the associations within sensitivity analyses using diagnostic codes based on published recommendations (18) and provides mechanistic evidence by using validated biomarkers to determine the potential influence of each locus on NAFLD development. Notably, using consensus recommendations to define NAFLD has resulted in 4,761 eligible cases whilst previous studies have classified only 704 individuals from within the UKB as NAFLD cases with the remainder misclassified as controls (14).

The limitations of this study include misclassification of individuals with alternative hepatic pathology on administrative records. The definition of cases based on administrative records also prevents any assessment of NAFLD severity and thus variants which contribute solely to progression of steatohepatitis, fibrosis or cirrhosis without promoting initial occurrence of steatosis, may not be identified. The binary case definition also offers lower power to detect associations than continuous traits. The results of our study do, however, support published literature with strong associations seen at established NAFLD-susceptibility loci. The GS-SFHS replication cohort only demonstrated significant association at the *GCKR* locus but none of the other known loci or the novel locus. This is likely to be due to the underpowered sample size with only 67 NAFLD cases compared to almost 5,000 analysed in the UKB cohort. The overall rate of NAFLD ascertained using the EHR-based approach was similar between the cohorts (1.2% in the UKB and 1.1% in the GS-SFHS cohorts) suggesting that the lack of replication may be determined by sample size rather than differences in clinical coding. Despite this, 5 of the 6 identified loci have been detected by earlier GWAS whilst the *APOE* locus has been investigated with significant results in candidate gene studies as well as reaching the threshold for replication in published GWAS summary association statistics suggesting that none of the 6 loci are false positives. Whilst the definition of NAFLD is based on consensus recommendations (18) it is possible that regional variation in recording prior to these recommendations has influenced documentation of NAFLD. Finally, the definition does not extend to more important clinical outcomes such as cirrhosis, hepatocellular carcinoma and cardiometabolic outcomes.

This paper supports the use of administrative data as a means to conduct research into NAFLD. Such approaches are likely to require large sample sizes but do overcome the need for invasive and costly recruitment and investigations.

In summary we have performed a GWAS of NAFLD using UKB data and identified six loci associated with NAFLD including rs429358-C. We have also demonstrated the feasibility of EHR-based NAFLD research without reliance on invasive investigations.

## Supporting information

Description of Supplementary Materials

## Data Availability

Raw data used in this manuscript is available from the UK Biobank and Generation Scotland. Both studies require an application from a verified researcher.

## List of Abbreviations

## Acknowledgements

Generation Scotland received core support from the Chief Scientist Office of the Scottish Government Health Directorates [CZD/16/6] and the Scottish Funding Council [HR03006] and is currently supported by the Wellcome Trust [216767/Z/19/Z]. Genotyping of the GS:SFHS samples was carried out by the Genetics Core Laboratory at the Edinburgh Clinical Research Facility, University of Edinburgh, Scotland and was funded by the Medical Research Council UK and the Wellcome Trust (Wellcome Trust Strategic Award “STratifying Resilience and Depression Longitudinally” (STRADL) Reference 104036/Z/14/Z).

This work was funded by a Medical Research Council Clinical Research Training Fellowship awarded to CJF (MR/T008008/1). ADB acknowledges funding from the Wellcome PhD training fellowship for clinicians (204979/Z/16/Z), the Edinburgh Clinical Academic Track (ECAT) programme. CH and JFW acknowledges support from the MRC Human Genetics Unit programme grant, “Quantitative traits in health and disease” (U. MC_UU_00007/10). NCH is supported by a Wellcome Trust Senior Research Fellowship in Clinical Science (ref. 219542/Z/19/Z). NLM is supported by the British Heart Foundation through a Senior Clinical Research Fellowship (FS/16/14/32023) and a Research Excellence Award (RE/18/5/34216). RKS is supported by the Wellcome Trust (grant 210752/Z/18/Z).

We acknowledge support from Dr Colin Fischbacher in selection of ICD10, ICD9 and Read codes.

## Author Contributions

CJF conducted the genome-wide association study and drafted the manuscript. CJF, ADB, PKJ, DWC, PRHJT and JFW developed the code required for data analysis. CJF, TMD and JFW assisted with data access requests. RP, EMH and JFW assisted with data storage and server maintenance. DJP, CH and AC contributed data from Generation Scotland. CJF, EMH, AS, SJW, CAS, TMD, RKS, NLM, CLMS and RP were involved in study design and conception. JAF, NCH, PR, NLM, RKS, SJW and EMH were involved in critical appraisal of the manuscript and placing the research in the biological context. ADB, AC, CH, JFW and AS were involved in critical appraisal of the manuscript in relation to GWAS methodology.

Authors names in bold designate shared co-first authorship.

## References

1. Younossi Z, Anstee QM, Marietti M, Hardy T, Henry L, Eslam M, et al. Global burden of NAFLD and NASH: Trends, predictions, risk factors and prevention. Nat Rev Gastro Hepat. 2018;15(1):11–20.

2. Dufour J-F, Caussy C, Loomba R. Combination therapy for non-alcoholic steatohepatitis: Rationale, opportunities and challenges. Gut. 2020;69(10):1877–1884.

3. Sookoian S, Pirola CJ. Genetic predisposition in nonalcoholic fatty liver disease. Clin Mol Hepat. 2017;23(1):1–12.

4. Romeo S, Kozlitina J, Xing C, Pertsemlidis A, Cox D, Pennacchio LA, et al. Genetic variation in PNPLA3 confers susceptibility to nonalcoholic fatty liver disease. Nat Genet. 2008;40(12):1461–1465.

5. Chalasani N, Guo X, Loomba R, Goodarzi MO, Haritunians T, Kwon S, et al. Genome-Wide Association Study Identifies Variants Associated with Histologic Features of Nonalcoholic Fatty Liver Disease. Gastroenterology. 2010;139(5):1567–1576.e6.

6. Kawaguchi T, Sumida Y, Umemura A, Matsuo K, Takahashi M, Takamura T, et al. Genetic Polymorphisms of the Human PNPLA3 Gene Are Strongly Associated with Severity of Non-Alcoholic Fatty Liver Disease in Japanese. PLoS ONE. 2012;7(6):1–10.

7. Adams LA, White SW, Marsh JA, Lye SJ, Connor KL, Maganga R, et al. Association between liver-specific gene polymorphisms and their expression levels with nonalcoholic fatty liver disease. Hepatology. 2013;57(2):590–600.

8. Kitamoto T, Kitamoto A, Yoneda M, Hyogo H, Ochi H, Nakamura T, et al. Genome-wide scan revealed that polymorphisms in the PNPLA3, SAMM50, and PARVB genes are associated with development and progression of nonalcoholic fatty liver disease in Japan. Hum Genet. 2013;132(7):783–792.

9. Wood KL, Miller MH, Dillon JF. Systematic review of genetic association studies involving histologically confirmed non-alcoholic fatty liver disease. BMJ Open Gastroenterol. 2015;2(1):1–19.

10. Wattacheril J, Lavine JE, Chalasani NP, Guo X, Kwon S, Schwimmer J, et al. Genome Wide Associations Related to Hepatic Histology in Nonalcoholic Fatty Liver Disease in Hispanic Boys. J Pediatri. 2017;190:1–16.

11. Chung GE, Lee Y, Yim JY, Choe EK, Kwak M-S, Yang JI, et al. Genetic Polymorphisms of PNPLA3 and SAMM50 Are Associated with Nonalcoholic Fatty Liver Disease in a Korean Population. Gut and Liver. 2018;12(3):316–323.

12. Namjou B, Lingren T, Huang Y, Parameswaran S, Cobb BL, Stanaway IB, et al. GWAS and enrichment analyses of non-alcoholic fatty liver disease identify new trait-associated genes and pathways across eMERGE Network. BMC Med. 2019;17(135):1–19.

13. Anstee QM, Darlay R, Cockell S, Meroni M, Govaere O, Tiniakos D, et al. Genome-wide association study of non-alcoholic fatty liver and steatohepatitis in a histologically characterised cohort. J Hepatol. 2020;73(3):505–515.

14. Emdin CA, Haas ME, Khera AV, Aragam K, Chaffin M, Klarin D, et al. A missense variant in Mitochondrial Amidoxime Reducing Component 1 gene and protection against liver disease. PLOS Genet. 2020;16(4):1–16.

15. Parisinos CA, Wilman HR, Thomas EL, Kelly M, Nicholls RC, McGonigle J, et al. Genome-wide and Mendelian randomisation studies of liver MRI yield insights into the pathogenesis of steatohepatitis. J Hepatol. 2020;73(2):241–251.

16. Park SL, Li Y, Sheng X, Hom V, Xia L, Zhao K, et al. Genome-Wide Association Study of Liver Fat: The Multiethnic Cohort Adiposity Phenotype Study. Hepatol Commun. 2020;4(8):1112–1123.

17. Yoshida K, Yokota K, Kutsuwada Y, Nakayama K, Watanabe K, Matsumoto A, et al. Genome-Wide Association Study of Lean Nonalcoholic Fatty Liver Disease Suggests Human Leukocyte Antigen as a Novel Candidate Locus. Hepatol Commun. 2020;4(8):1124–1135.

18. Hagström H, Adams LA, Allen AM, Byrne CD, Chang Y, Grønbæk H, et al. Administrative coding in electronic health care record-based research of NAFLD: An expert panel consensus statement. Hepatology. 2021;Online ahead of print.

19. UK Biobank. UK Biobank: Protocol for a large-scale prospective epidemiological resource. Stockport, UK: UK Biobank; 2007 pp. 1–112. Report No.: UKBB-PROT-09-06.

20. Elliott P, Peakman TC, UK Biobank. The UK Biobank sample handling and storage protocol for the collection, processing and archiving of human blood and urine. Int J Epidemiol. 2008;37(2):234–244.

21. Sudlow C, Gallacher J, Allen N, Beral V, Burton P, Danesh J, et al. UK biobank: An open access resource for identifying the causes of a wide range of complex diseases of middle and old age. PLoS Med. 2015;12(3):1–10.

22. Smith BH, Campbell H, Blackwood D, Connell J, Connor M, Deary IJ, et al. Generation Scotland: The Scottish Family Health Study; a new resource for researching genes and heritability. BMC Med Genet. 2006;7(74):1–9.

23. Smith BH, Campbell A, Linksted P, Fitzpatrick B, Jackson C, Kerr SM, et al. Cohort Profile: Generation Scotland: Scottish Family Health Study (GS:SFHS). The study, its participants and their potential for genetic research on health and illness. Int J Epidemiol. 2013;42(3):689–700.

24. Manichaikul A, Mychaleckyj JC, Rich SS, Daly K, Sale M, Chen W-M. Robust relationship inference in genome-wide association studies. Bioinformatics. 2010;26(22):2867–2873.

25. Watanabe K, Taskesen E, van Bochoven A, Posthuma D. Functional mapping and annotation of genetic associations with FUMA. Nat Commun. 2017;8(1):1–11.

26. Bulik-Sullivan BK, Loh P-R, Finucane HK, Ripke S, Yang J, Patterson N, et al. LD Score regression distinguishes confounding from polygenicity in genome-wide association studies. Nat Genet. 2015;47(3):291–295.

27. Yoneda M, Mawatari H, Fujita K, Iida H, Yonemitsu K, Kato S, et al. High-sensitivity C-reactive protein is an independent clinical feature of nonalcoholic steatohepatitis (NASH) and also of the severity of fibrosis in NASH. J Gastroenterol. 2007;42(7):573–582.

28. Verma S, Jensen D, Hart J, Mohanty SR. Predictive value of ALT levels for non-alcoholic steatohepatitis (NASH) and advanced fibrosis in non-alcoholic fatty liver disease (NAFLD). Liver Int. 2013;33(9):1398–1405.

29. Angulo P, Hui JM, Marchesini G, Bugianesi E, George J, Farrell GC, et al. The NAFLD fibrosis score: A noninvasive system that identifies liver fibrosis in patients with NAFLD. Hepatology. 2007;45(4):846–854.

30. Vallet-Pichard A, Mallet V, Nalpas B, Verkarre V, Nalpas A, Dhalluin-Venier V, et al. FIB-4: An inexpensive and accurate marker of fibrosis in HCV infection. Comparison with liver biopsy and fibrotest. Hepatology. 2007;46(1):32–36.

31. Willer CJ, Li Y, Abecasis GR. METAL: Fast and efficient meta-analysis of genomewide association scans. Bioinformatics. 2010;26(17):2190–2191.

32. Begum F, Ghosh D, Tseng GC, Feingold E. Comprehensive literature review and statistical considerations for GWAS meta-analysis. Nucleic Acids Res. 2012;40(9):3777– 3784.

33. Marchini J, Howie B. Genotype imputation for genome-wide association studies. Nat Rev Genet. 2010;11(7):499–511.

34. R Core Team. R: A Language and Environment for Statistical Computing. Vienna, Austria: R Foundation for Statistical Computing; 2017.

35. Little J, Higgins JPT, Ioannidis JPA, Moher D, Gagnon F, Elm E von, et al. STrengthening the REporting of Genetic Association Studies (STREGA) of the STROBE Statement. PLOS Med. 2009;6(2):0151–0163.

36. Liu Y-L, Reeves HL, Burt AD, Tiniakos D, McPherson S, Leathart JBS, et al. TM6SF2 rs58542926 influences hepatic fibrosis progression in patients with non-alcoholic fatty liver disease. Nat Commun. 2014;5(4309):1–6.

37. Yang J, Weedon MN, Purcell S, Lettre G, Estrada K, Willer CJ, et al. Genomic inflation factors under polygenic inheritance. Eur J Hum Genet. 2011;19(7):807–812.

38. Kern S, Mehlig K, Kern J, Zetterberg H, Thelle D, Skoog I, et al. The Distribution of Apolipoprotein E Genotype Over The Adult Lifespan and in Relation to Country of Birth. Am J Epidemiol. 2015;181(3):214–217.

39. Bennet AM, Angelantonio ED, Ye Z, Wensley F, Dahlin A, Ahlbom A, et al. Association of Apolipoprotein E Genotypes With Lipid Levels and Coronary Risk. JAMA. 2007;298(11):1300–1311.

40. Palmer ND, Kahali B, Kuppa A, Chen Y, Du X, Feitosa MF, et al. Allele Specific Variation at APOE Increases Non-alcoholic Fatty Liver Disease and Obesity but Decreases Risk of Alzheimer’s Disease and Myocardial Infarction. Hum Mol Genet. 2021;[In Press].

41. Yang MH, Son HJ, Sung JD, Choi YH, Koh KC, Yoo BC, et al. The relationship between apolipoprotein E polymorphism, lipoprotein (a) and fatty liver disease. Hepato-Gastroenterol. 2005;52(66):1832–1835.

42. Sazci A, Akpinar G, Aygun C, Ergul E, Senturk O, Hulagu S. Association of Apolipoprotein E Polymorphisms in Patients with Non-Alcoholic Steatohepatitis. Digest Dis Sci. 2008;53(12):3218–3224.

43. De Feo E, Cefalo C, Arzani D, Amore R, Landolfi R, Grieco A, et al. A caseControl study on the effects of the apolipoprotein E genotypes in nonalcoholic fatty liver disease. Mol Biol Rep. 2012;39(7):7381–7388.

44. Demirag MD, Onen HI, Karaoguz MY, Dogan I, Karakan T, Ekmekci A, et al. Apolipoprotein E Gene Polymorphism in Nonalcoholic Fatty Liver Disease. Digest Dis Sci. 2007;52(12):3399–3403.

45. Lee D-M, Lee S-O, Mun B-S, Ahn H-S, Park H-Y, Lee H-S, et al. [Relation of apolipoprotein E polymorphism to clinically diagnosed fatty liver disease]. Taehan Kan Hakhoe Chi = Korean J Hep. 2002;8(4):355–362.

46. van den Berg EH, Corsetti JP, Bakker SJL, Dullaart RPF. Plasma ApoE elevations are associated with NAFLD: The PREVEND Study. PLoS ONE. 2019;14(8):1–15.

47. Stachowska E, Maciejewska D, Ossowski P, Drozd A, Ryterska K, Banaszczak M, et al. Apolipoprotein E4 allele is associated with substantial changes in the plasma lipids and hyaluronic acid content in patients with nonalcoholic fatty liver disease. J Physiol Pharmacol. 2013;64(6):711–717.

48. Riches FM, Watts GF, van Bockxmeer FM, Hua J, Song S, Humphries SE, et al. Apolipoprotein B signal peptide and apolipoprotein E genotypes as determinants of the hepatic secretion of VLDL apoB in obese men. J Lipid Res. 1998;39(9):1752–1758.

49. Schierwagen R, Maybüchen L, Zimmer S, Hittatiya K, Bäck C, Klein S, et al. Seven weeks of Western diet in apolipoprotein-E-deficient mice induce metabolic syndrome and non-alcoholic steatohepatitis with liver fibrosis. Sci Rep. 2015;5(12931):1–14.

50. Kypreos KE, van Dijk KW, van der Zee A, Havekes LM, Zannis VI. Domains of Apolipoprotein E Contributing to Triglyceride and Cholesterol Homeostasis in Vivo: Carboxyl-Terminal Region 203299 Promotes Hepatic Very-low-density-lipoprotein-triglyceride Secretion. J Biol Chem. 2001;276(23):19778–19786.

