## Supplementary material for "Genome-Wide Association Study of Non-Alcoholic Fatty Liver Disease Identifies Association with Apolipoprotein E": Table 1

**Loci associated with non-alcoholic fatty liver disease in the UK Biobank Cohort**

| RSID | Chr:Pos | Maj/Min | MAF | OR  (95% CI) | P Value | Consequence (Gene) | Hypothesised Functional Gene |
| --- | --- | --- | --- | --- | --- | --- | --- |
| rs2642442 | 1:220973563 | C/T | 0.317 | 1.15  (1.10-1.20) | 7.67e-10 | Intron (MARC1) | MARC1 |
| rs1260326 | 2:27730940 | T/C | 0.392 | 0.87  (0.84-0.91) | 2.54e-11 | Missense (GCKR) | GCKR |
| rs17321515 | 8:126486409 | A/G | 0.476 | 0.86  (0.82-0.89) | 1.81e-13 | Intergenic (TRIB1) | TRIB1 |
| rs73001065 | 19:19460541 | G/C | 0.071 | 1.41  (1.32-1.51) | 1.08e-24 | Intron (MAU2) | TM6SF2 |
| rs429358 | 19:45411941 | T/C | 0.156 | 0.82  (0.77-0.87) | 2.17e-11 | Missense (APOE) | APOE |
| rs3747207 | 22:44324855 | G/A | 0.215 | 1.45  (1.38-1.51) | 6.74e-60 | Intron (PNPLA3) | PNPLA3 |
| Chr:Pos: Chromosome:Position, Maj/Min: Major allele/ Minor allele, MAF: Minor allele frequency, P Value: P value using allelic model, OR (95% CI): Odds ratio with 95% confidence interval. Functional Role is based on assessment of published literature. Chromosome and position based on Genome Reference Consortium Human Build 37. Effect allele is the minor allele. | | | | | | | |
