## Supplementary material for "Genome-Wide Association Study of Non-Alcoholic Fatty Liver Disease Identifies Association with Apolipoprotein E": Table 2

**Association of identified loci with non-alcoholic fatty liver disease in the replication cohort**

|  |  |  | GS-SFHS | | | Namjou et al. | | Anstee et al. | | Meta-analysis |  |
| --- | --- | --- | --- | --- | --- | --- | --- | --- | --- | --- | --- |
| RSID | Chr:Pos | Maj/Min | MAF | OR  (95% CI) | P Value | OR*^1^* | P Value | OR  (95% CI) | P Value | P Value | Gene |
| rs2642442 | 1:220973563 | C/T | 0.311 | 1.06  (0.73-1.54) | 0.551 | 1.19 | 5.96e-03 | 1.16  (1.06-1.26) | 9.70e-04 | 5.83e-12 | MARC1 |
| rs1260326 | 2:27730940 | T/C | 0.387 | 0.73  (0.52-1.03) | 0.00737 | 0.90 | 7.34e-02 | 0.78  (0.73-0.84) | 1.06e-10 | 3.08e-15 | GCKR |
| rs17321515 | 8:126486409 | A/G | 0.478 | 0.94  (0.67-1.32) | 0.913 | 0.80 | 1.08e-04 | 0.86  (0.79-0.93) | 1.99e-04 | 1.24e-16 | TRIB1 |
| rs73001065 | 19:19460541 | G/C | 0.065 | 0.79  (0.37-1.69) | 0.591 | 1.30 | 1.19e-02 | 1.58  (1.37-1.82) | 1.59e-10 | 7.51e-30 | TM6SF2 |
| rs429358 | 19:45411941 | T/C | 0.162 | 0.70  (0.41-1.18) | 0.126 | 0.81 | 9.57e-03 | 0.85  (0.77-0.95) | 4.16e-03 | 3.42e-13 | APOE |
| rs3747207 | 22:44324855 | G/A | 0.194 | 1.37  (0.92-2.03) | 0.142 | 1.78 | 2.63e-20 | 1.83  (1.68-1.98) | 2.58e-49 | 1.67e-87 | PNPLA3 |
| *^1^*The summary association statistics for Namjou et al. did not provide a confidence interval or standard error for the odds ratio.  Chr:Pos: Chromosome:Position, Maj/Min: Major allele/ Minor allele, MAF: Minor allele frequency, P Value: P value using allelic model, OR (95% CI): Odds ratio with 95% confidence interval. Chromosome and position based on Genome Reference Consortium Human Build 37. | | | | | | | | | | | |
