## Supplementary material for "Genome-Wide Association Study of Non-Alcoholic Fatty Liver Disease Identifies Association with Apolipoprotein E": Description of Supplementary Materials

### Supplementary Materials 1 - Diagnostic Codes

- Tables of ICD9, ICD10, UK Biobank, Read V2 and Read V3 codes
  - Non-alcoholic fatty liver disease (NAFLD)
  - Other hepatobiliary pathology for exclusions

### Supplementary Materials 2 - Significant GWAS Results*

- xlsx file with main significant SNPs for main GWAS and subgroup analyses, sheets of xlsx file as follows:
  - “NAFLD versus Controls”: SNPtest output for all SNPs reaching genome-wide significance (P<5*10^-8^) in main GWAS (6 Main GWAS SNPs)
  - “Lead SNPs”: SNPtest output for the 6 SNPs from the main GWAS identified as lead SNPs
  - “Differentials Excluded”: SNPtest output for all 6 Main GWAS SNPs in GWAS comparing NAFLD to controls after exclusion of alternative hepatic pathology
  - “NAFLD vs Controls (BMI)”: SNPtest output for all 6 Main GWAS SNPs in GWAS adjusted for body mass index
  - “NAFLD vs Controls (Alcohol)”: SNPtest output for all 6 Main GWAS SNPs in GWAS adjusted for body mass index and alcohol intake
  - “Replication”: Results of the Generation Scotland: Scottish Family Health Study data at each of the NAFLD-susceptibility loci
  - “Namjou et al”: Published GWA summary association statistics from Namjou et al for the 6 lead SNPs in the UK Biobank study
  - “Anstee et al”: Published GWA summary association statistics from Anstee et al for the 6 lead SNPs in the UK Biobank study
  - “GWAS Meta-analysis”: METAL GWAS output for the 6 lead SNPs

### Supplementary Materials 3A - FUMA Parameters and Data Dictionary*

- Data dictionary for each file in the main FUMA results
- List of parameters passed to the Functional Mapping and Annotation of GWAS results

### Supplementary Materials 3B - FUMA Results*

- FUMA results in xlsx file (each sheet of the file is a specific FUMA output file as described in 3A)

### Supplementary Materials 4 - Conditional Analyses Results*

- Results for each of the conditional analyses in a separate sheet of an xlsx file
  - SNPtest output for variants with genome-wide significance after conditional analyses shown (loci boundaries available in FUMA results - see 3B)

### Supplementary Materials 5 - Analysis of Apolipoprotein E Genotype Associations

- Methods describing analyses undertaken using apolipoprotein E genotype
- Table of APOE genotype status associations with NAFLD
- Odds ratio plots for association of apoliprotein E genotype with NAFLD

### Supplementary Materials 6 - Additional Figures

- Supplementary Tables:
  - Supplementary Table 1: Description of Published Case-Control GWAS Studies
  - Supplementary Table 2: Replication of Published Case-Control GWAS for NAFLD
  - Supplementary Table 3: Baseline Characteristics of the UK Biobank NAFLD GWAS cohort
- Lattice plots for serum biochemistry linear regressions:
  - Supplementary Figure 1: Impact of NAFLD-susceptibility alleles on other lipid particles (apolipoprotein A and B and lipoprotein A)
  - Supplementary Figure 2: Impact of NAFLD-susceptibility alleles on liver enzymes (alanine aminotransferase, aspartate aminotransferase, alkaline phosphatase and gamma glutamyltransferase)
  - Supplementary Figure 3: Impact of NAFLD-susceptibility alleles on glycaemic control (glucose and glycated haemoglobin)
  - Supplementary Figure 4: Impact of NAFLD-susceptibility alleles on C-reactive protein
  - Supplementary Figure 5: Impact of NAFLD-susceptibility alleles on anthropometric features (body mass index, waist circumference, hip circumference and waist:hip ratio)
- Lattice plot comparing main GWAS results with subgroup analyses:
  - Supplementary Figure 6: Comparison of effect sizes at NAFLD-susceptibility alleles between the main GWAS and the “Differentials Excluded” subgroup analysis
- GWA Meta-analysis figures:
  - Supplementary Figure 7: Comparison of effect sizes between the UK Biobank GWAS and other published NAFLD GWAS summary association statistics at the 6 identified lead SNPs
  - Supplementary Figure 8: Updated Manhattan plot using GWAS meta-analysis data
- Quantile-Quantile Plot
  - Supplementary Figure 9: Quantile-Quantile plot for the UK Biobank Non-Alcoholic Fatty Liver Disease GWAS

### Supplementary Materials 7 - Biomarker Regression Results*

- Numeric results from each of the biomarker and anthropometric features lattice plots covering linear regression coefficients:
  - xlsx file which contains one sheet per trait (total cholesterol, low-density lipoprotein, triglycerides, high-density lipoprotein, apolipoprotein A, apolipoprotein B, lipoprotein A, alkaline phosphatase, alanine aminotransferase, aspartate aminotransferase, gamma glutamyltransferase, glucose, glycated haemoglobin, C-reactive protein, body mass index, hip circumference, waist circumference, wasit:hip ratio)

### Supplementary Materials 8 – STREGA Checklist

- STrengthening the REporting of Genetic Association Studies checklist

***Data made available through download interface hosted at <https://argoshare.is.ed.ac.uk/connect/#/apps/532/access>**

**Supplementary material including output for all of the significant SNPs has been made available through this interface as the files run to several thousands of lines and are too large to attach with the remaining Supplementary Materials.**
