## Supplementary Materials 1 for "Genome-Wide Association Study of Non-Alcoholic Fatty Liver Disease Identifies Association with Apolipoprotein E"

Supplementary Materials 2, 3A, 3B, 4 and 7

Supplementary material including output for all of the significant SNPs has been made available through a separate interface as the files run to several thousands of lines and are too large to attach with the remaining Supplementary Materials.

The files are available for inspection at:

<https://argoshare.is.ed.ac.uk/connect/#/apps/532/access>
