## Supplementary Materials 2, 3A, 3B, 4 and 7 for "Genome-Wide Association Study of Non-Alcoholic Fatty Liver Disease Identifies Association with Apolipoprotein E"

Supplementary Material - Apolipoprotein E Analysis

Rs429358 is a common variant within apolipoprotein E (APOE) which in combination with rs7412 governs determination of the three main alleles of APOE (ϵ3, ϵ4 and ϵ2)1. The main apolipoprotein E genotypes (ϵ3ϵ3, ϵ2ϵ3, ϵ3ϵ4, ϵ2ϵ4, ϵ4ϵ4 and ϵ2ϵ2) have previously been investigated for association with NAFLD with some studies showing no difference whilst others have showed elevated risk with the common ϵ3 alleles^2–8^.

As rs429358 was identified as a risk factor for non-alcoholic fatty liver disease (NAFLD) within the main GWAS, an analysis of the genotype status and risk of NAFLD was undertaken.

### Methods

Both rs429358 and rs7412 were directly called within the genotyping arrays meaning that allelic dosages were available in integer format. Genotype dosage was retrieved from the files using QCTOOLS version 2.0.6^9^.

Genotype was determined based on number of alternate alleles at each SNP (Table 6-I).

As the genotype data was unphased it was not possible to determine whether rs7412 TC and rs429358 TC corresponded to a TT and CC haplotype or two CT haplotypes and as such it is not certain whether the patient was of a $\epsilon2\epsilon4$ or a $\epsilon1\epsilon3$ genotype. However, given the extreme rarity of the $\epsilon1$ allele it was determined that patients who were heterozygotes at both SNPs should be considered as $\epsilon2\epsilon4$ genotypes. The other $\epsilon1$ allelic carriers were excluded from the analysis based on rarity of the genotype (see Results section).

Unadjusted association between APOE genotype and NAFLD was tested with $\chi2$ test. The association was then evaluated in a logistic regression with the 6 included APOE genotypes. Each of these logistic regressions was adjusted for age and sex and the genetic principal components within the main GWAS model.

### Results

A total of 377,998 individuals had sufficient data available to calculate APOE genotype. As expected, the most common genotype was $\epsilon3\epsilon3$ (219,869) whilst there were a total of 19 individuals who were either $\epsilon1\epsilon4$ or $\epsilon1\epsilon2$ who were excluded (there were no $\epsilon1\epsilon1$ homozygotes). The distribution of individuals by genotype is shown in Table 6-II.

APOE genotype was highly significantly associated with NAFLD status (χ^2^=2.49*10^-8^), see Supplementary Figure 6-I). This association appears to be driven by decreased overall

risk for every $\epsilon4$ allele and with a trend towards lower risk with the $\epsilon2$ allele. The association was evident on logistic regression for any of the three $\epsilon4$ allele-carrying genotypes (odds ratio [OR] and 95% confidence interval [95%CI] for ϵ4ϵ4: 0.64 [0.50-0.79]; for ϵ3ϵ4: 0.84 [0.78-0.90]; and for ϵ2ϵ4 0.74 [0.60-0.90]). The other ϵ2 allele-carrying genotypes were not associated with change in NAFLD risk (ϵ2ϵ2 OR 0.76 [0.50-1.12]; and ϵ2ϵ3 1.02 [0.93-1.11]).

The effect sizes for each of the apolipoprotein E genotypes are shown in an odds ratio plot in the main paper.

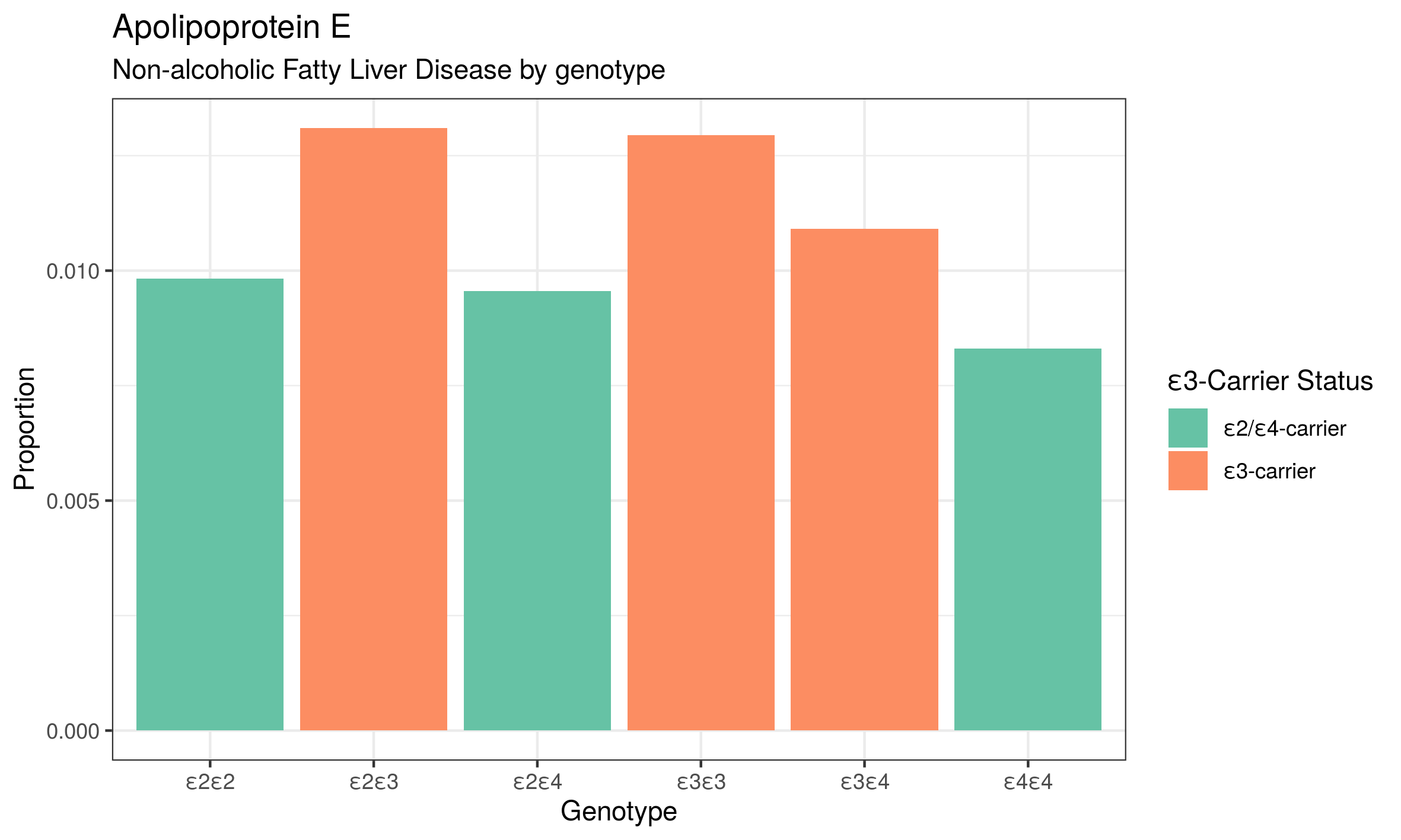

Supplementary Figure 6-I: Bar plot demonstrating proportion of NAFLD cases by each apolipoprotein E genotype.

**References**

1. Kern, S. *et al.* The Distribution of Apolipoprotein E Genotype Over The Adult Lifespan and in Relation to Country of Birth. *American Journal of Epidemiology* **181**, 214–217 (2015).

2. Lee, D.-M. *et al.* [Relation of apolipoprotein E polymorphism to clinically diagnosed fatty liver disease]. *Taehan Kan Hakhoe Chi = The Korean Journal of Hepatology* **8**, 355–362 (2002).

3. Yang, M. H. *et al.* The relationship between apolipoprotein E polymorphism, lipoprotein (a) and fatty liver disease. *Hepato-Gastroenterology* **52**, 1832–1835

4. Demirag, M. D. *et al.* Apolipoprotein E Gene Polymorphism in Nonalcoholic Fatty Liver Disease. *Digestive Diseases and Sciences* **52**, 3399–3403 (2007).

5. Sazci, A. *et al.* Association of Apolipoprotein E Polymorphisms in Patients with Non-Alcoholic Steatohepatitis. *Digestive Diseases and Sciences* **53**, 3218–3224 (2008).

6. Stachowska, E. *et al.* Apolipoprotein E4 allele is associated with substantial changes in the plasma lipids and hyaluronic acid content in patients with nonalcoholic fatty liver disease. *Journal of Physiology and Pharmacology: An Official Journal of the Polish Physiological Society* **64**, 711–717 (2013).

7. De Feo, E. *et al.* A caseControl study on the effects of the apolipoprotein E genotypes in nonalcoholic fatty liver disease. *Molecular Biology Reports* **39**, 7381–7388 (2012).

8. van den Berg, E. H., Corsetti, J. P., Bakker, S. J. L. & Dullaart, R. P. F. Plasma ApoE elevations are associated with NAFLD: The PREVEND Study. *PLoS ONE* **14**, (2019).

9. Marchini, J. & Howie, B. Genotype imputation for genome-wide association studies. *Nature Reviews Genetics* **11**, 499–511 (2010).
