## Supplementary Materials 5 for "Genome-Wide Association Study of Non-Alcoholic Fatty Liver Disease Identifies Association with Apolipoprotein E"

Supplementary Material 6 - Additional Figures

1. Supplementary Table 1: Description of Published Case-Control GWAS Studies
2. Supplementary Table 2: Replication of Published Case-Control GWAS for NAFLD
3. Supplementary Table 3: Baseline Characteristics of the UK Biobank NAFLD GWAS cohort
4. Supplementary Figure 1: Association with Serum Lipid Particles
5. Supplementary Figure 2: Association with Serum Liver Enzymes
6. Supplementary Figure 3: Association with Serum Glucose and HbA1c
7. Supplementary Figure 4: Association with Serum C-Reactive Protein
8. Supplementary Figure 5: Association with Anthropometric Features
9. Supplementary Figure 6: Comparison of effect sizes between main GWAS and subgroup analyses (differentials excluded, adjusted for BMI and adjusted for BMI and alcohol intake)
10. Supplementary Figure 7: Comparison of effect sizes between the UK Biobank GWAS and other published NAFLD GWAS summary association statistics at the 6 identified lead SNPs
11. Supplementary Figure 8: Updated Manhattan plot using GWAS meta-analysis data
12. Supplementary Figure 9: Quantile-Quantile Plot for the UK Biobank NAFLD GWAS showing observed quantiles of variant effects as a function of expected effect sizes from a normal distribution

| Supplementary Table 1: Description of Published Case-Control GWAS for NAFLD | | | | |
| --- | --- | --- | --- | --- |
| Study First Author | Year | Cases | Controls | Diagnosis of NAFLD |
| Anstee | 2020 | 1483 | 17781 | Histological |
| Emdin*^1^* | 2020 | 704 | 404865 | Electronic Health Records |
| Yoshida | 2020 | 275 | 1411 | Radiological |
| Namjou | 2019 | 1106 | 8571 | Electronic Health Records |
| Chung | 2018 | 1593 | 2816 | Radiological |
| Adams | 2013 | 126 | 802 | Radiological |
| Kitamoto | 2013 | 392 | 934 | Histological |
| Kawaguchi | 2012 | 539 | 932 | Histological |
| *^1^*Emdin et al performed a GWAS of all-cause cirrhosis before assessing the relationship of just rs2642438 with non-alcoholic fatty liver disease (NAFLD). | | | | |

References listed in main manuscript.

| Supplementary Table 2: Replication of Published Case-Control GWAS for NAFLD | | | | | | |
| --- | --- | --- | --- | --- | --- | --- |
| Chromosome | Hypothesised Gene | Studies Reporting Gene | SNP Assessed in UK Biobank | Odds Ratio (95% CI) | P Value | Replicated? |
| 1 | MARC1 | Emdin 2020 | rs2642442 | 1.15  (1.10-1.20) | 7.67e-10 | Yes |
| 2 | GCKR | Anstee 2020 | rs1260326 | 0.87  (0.84-0.91) | 2.54e-11 | Yes |
| 4 | HSD17B13 | Anstee 2020 | rs13118664 | 0.94  (0.92-0.96) | 7.41e-03 | Yes |
| 19 | TM6SF2 | Chung 2018  Anstee 2020 | rs73001065 | 1.41  (1.32-1.51) | 1.08e-24 | Yes |
| 22 | PNPLA3 | Kawaguchi 2012  Chung 2013  Kitamoto 2013  Namjou 2019  Anstee 2020  Yoshida 2020 | rs3747207 | 1.45  (1.38-1.51) | 6.74e-60 | Yes |
| CI: Confidence Interval. Replication considered at P<0.0083 based on Bonferroni Correction | | | | | | |

| Supplementary Table 3: Baseline Characteristics of the UK Biobank NAFLD GWAS cohort | | | | |
| --- | --- | --- | --- | --- |
|  |  | NAFLD | Controls | p |
| Sex | Female | 2332 (49.0) | 200563 (53.7) | <0.001 |
|  | Male | 2429 (51.0) | 172664 (46.3) |  |
| Age at Recruitment | Mean (SD) | 57.4 (7.6) | 56.9 (7.9) | <0.001 |
| Weight (kg) | Mean (SD) | 89.2 (17.4) | 78.2 (15.8) | <0.001 |
| BMI (kg/m^2^) | Mean (SD) | 31.4 (5.4) | 27.4 (4.7) | <0.001 |
| Systolic Blood Pressure (mmHg) | Mean (SD) | 141.5 (17.5) | 138.3 (18.6) | <0.001 |
| Diastolic Blood Pressure (mmHg) | Mean (SD) | 84.9 (10.2) | 82.3 (10.1) | <0.001 |
| Alanine Aminotransferase (U/L) | Mean (SD) | 38.7 (26.2) | 23.4 (13.8) | <0.001 |
| Aspartate Aminotransferase (U/L) | Mean (SD) | 35.3 (21.9) | 26.1 (10.3) | <0.001 |
| Alkaline Phosphatase (U/L) | Mean (SD) | 95.4 (37.5) | 83.6 (26.4) | <0.001 |
| Gamma Glutamyltransferase (U/L) | Mean (SD) | 80.4 (96.9) | 37.0 (40.6) | <0.001 |
| Cholesterol (mmol/L) | Mean (SD) | 5.5 (1.3) | 5.7 (1.1) | <0.001 |
| Triglycerides (mmol/L) | Mean (SD) | 2.3 (1.3) | 1.8 (1.0) | <0.001 |
| Low-Density Lipoprotein (mmol/L) | Mean (SD) | 3.4 (1.0) | 3.6 (0.9) | <0.001 |
| High-Density Lipoprotein (mmol/L) | Mean (SD) | 1.3 (0.3) | 1.5 (0.4) | <0.001 |
| Apolipoprotein A (g/L) | Mean (SD) | 1.4 (0.3) | 1.5 (0.3) | <0.001 |
| Apolipoprotein B (g/L) | Mean (SD) | 1.0 (0.3) | 1.0 (0.2) | 0.868 |
| Lipoprotein A (nmol/l) | Mean (SD) | 42.0 (48.5) | 44.1 (49.5) | 0.010 |
| C-Reactive Protein (mg/L) | Mean (SD) | 4.2 (5.6) | 2.6 (4.3) | <0.001 |
| Glucose (mmol/L) | Mean (SD) | 5.7 (2.1) | 5.1 (1.2) | <0.001 |
| Glycated Haemoglobin (HbA1c; mmol/mol) | Mean (SD) | 40.0 (13.0) | 35.9 (6.4) | <0.001 |
| All Diabetes*^1^* | Yes | 1567 (32.9) | 28830 (7.7) | <0.001 |
|  | No | 3194 (67.1) | 344397 (92.3) |  |
| Type 2 Diabetes | Yes | 1520 (31.9) | 24796 (6.6) | <0.001 |
|  | No | 3241 (68.1) | 348431 (93.4) |  |
| *^1^*'All Diabetes' includes Type 1, Type 2, other types and unspecified diabetes mellitus. | | | | |
| Categorical data tested with Chi-squared test. Continuous data tested with Welch two-sample T-test. | | | | |

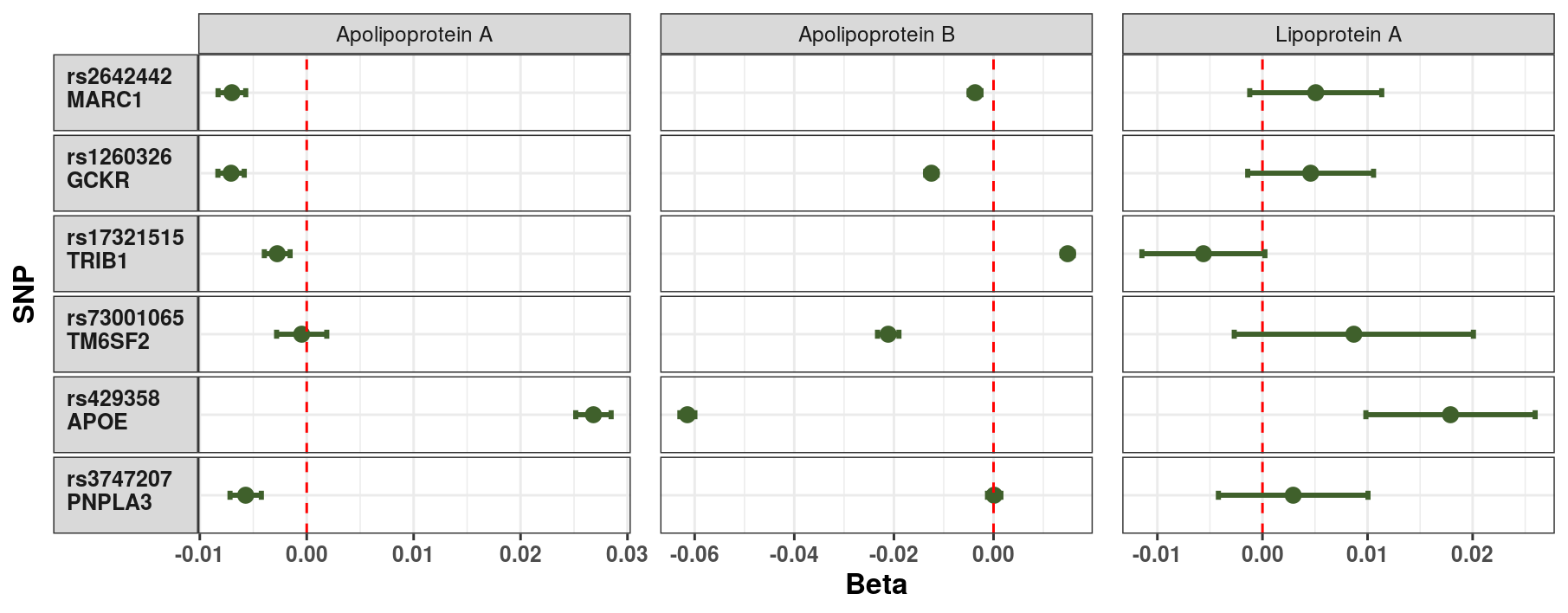

Supplementary Figure 1: Impact of each NAFLD-promoting allele on the measured serum lipid particles. Each point represents the beta-coefficient from an age- and sex-adjusted linear regression and the error bar represents the 95% confidence interval. Apo A - Apolipoprotein A; Apo B - Apolipoprotein B; Lipo A - Lipoprotein A. Lipoprotein A was log-transformed prior to analysis.

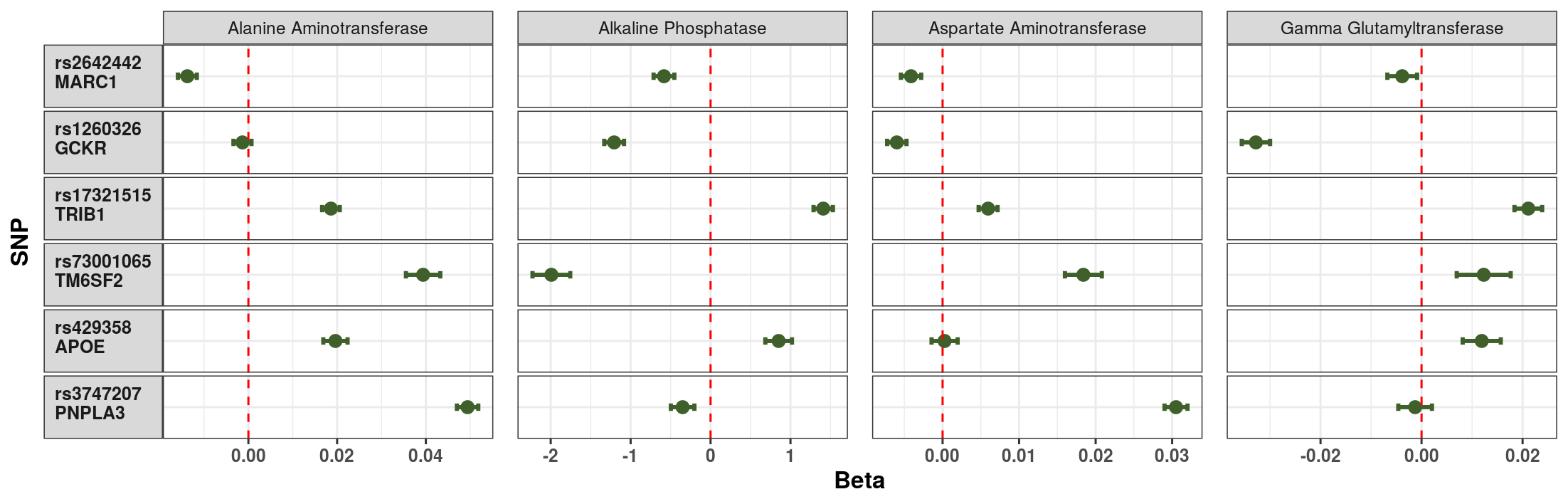

Supplementary Figure 2: Impact of each NAFLD-promoting allele on the measured serum liver enzymes. Each point represents the beta-coefficient from an age- and sex-adjusted linear regression and the error bar represents the 95% confidence interval. Alk P - Alkaline Phosphatase; ALT - Alanine Aminotransferase; AST - Aspartate Aminotransferase; GGT - Gamma Glutamyltransferase. Each of the serum biomarkers was log-transformed prior to analysis.

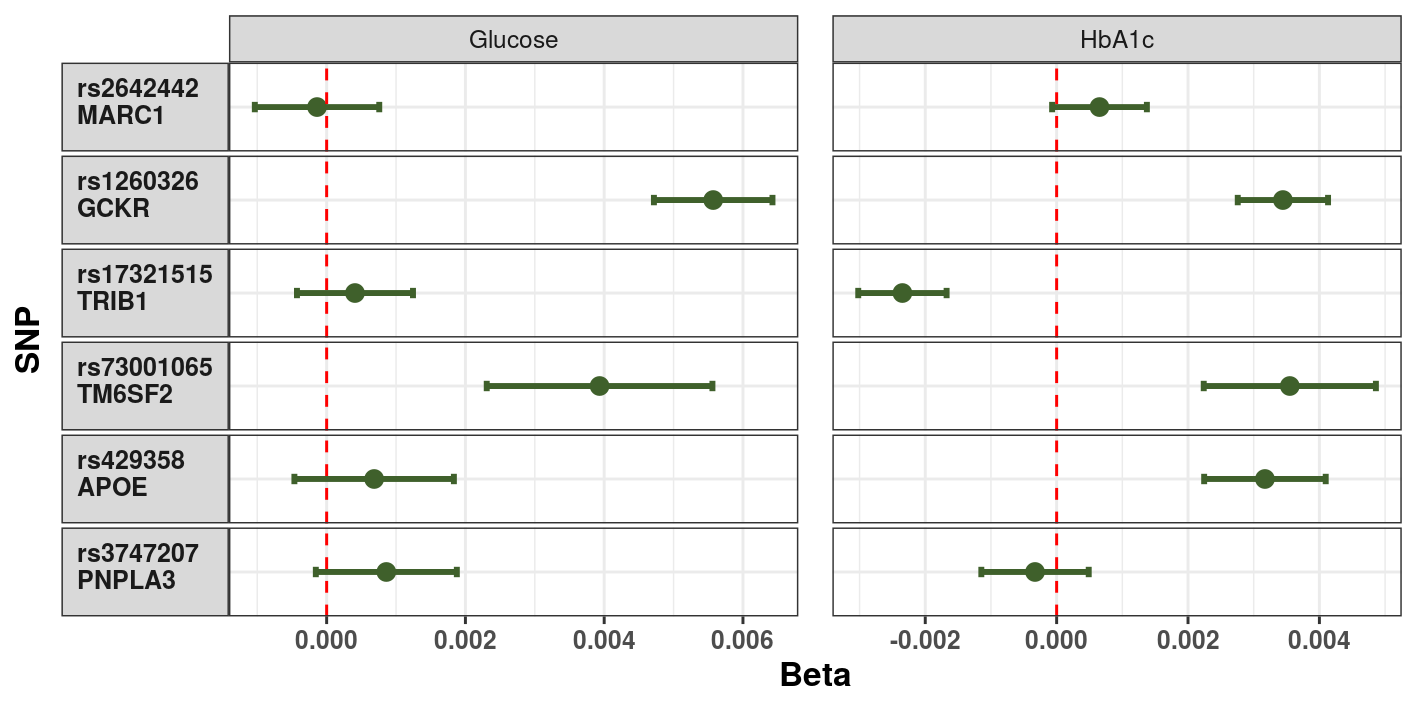

Supplementary Figure 3: Impact of each NAFLD-promoting allele on the measured serum glucose and glycated haemoglobin. Each point represents the beta-coefficient from an age- and sex-adjusted linear regression and the error bar represents the 95% confidence interval. HbA1c - glycated haemoglobin. Both glucose and HbA1c were log-transformed prior to analysis.

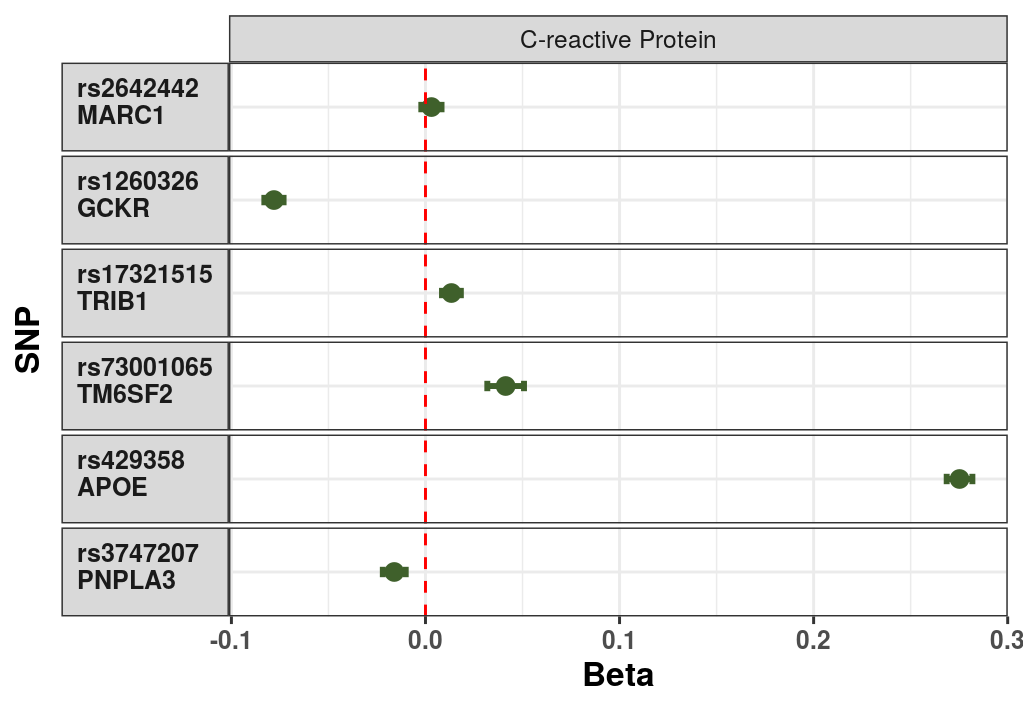

Supplementary Figure 4: Impact of each NAFLD-promoting allele on the measured serum C-reactive protein. Each point represents the beta-coefficient from an age- and sex-adjusted linear regression and the error bar represents the 95% confidence interval. CRP - C-reactive protein. CRP was log-transformed prior to analysis.

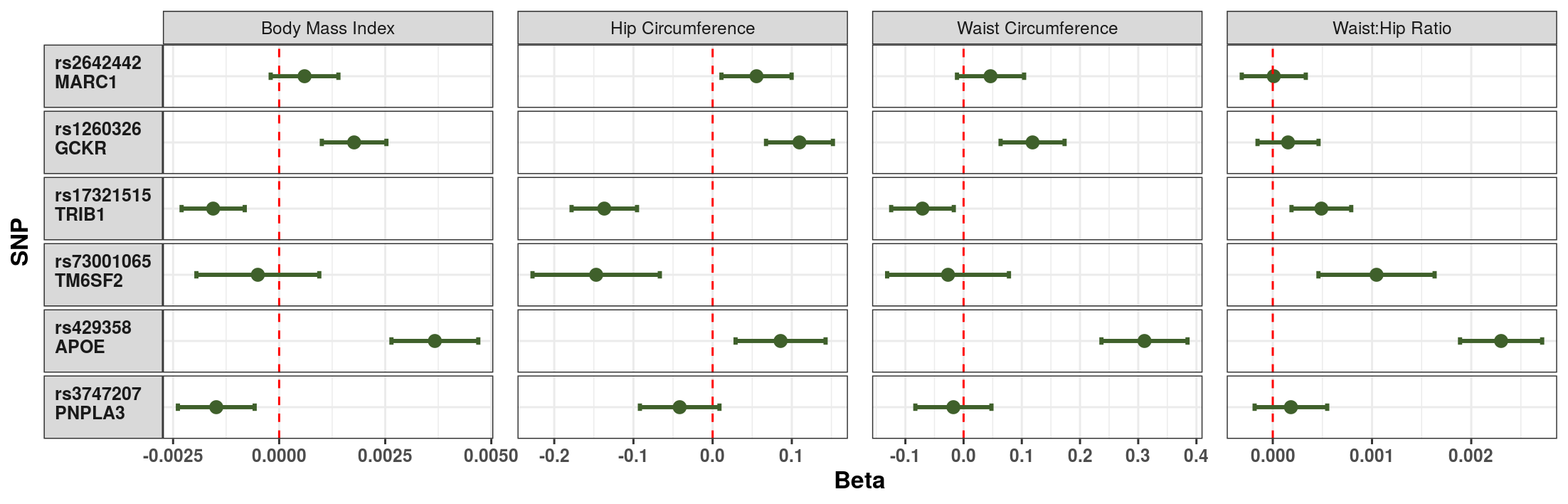

Supplementary Figure 5: Impact of each NAFLD-promoting allele on anthropometric features. Each point represents the beta-coefficient from an age- and sex-adjusted linear regression and the error bar represents the 95% confidence interval.

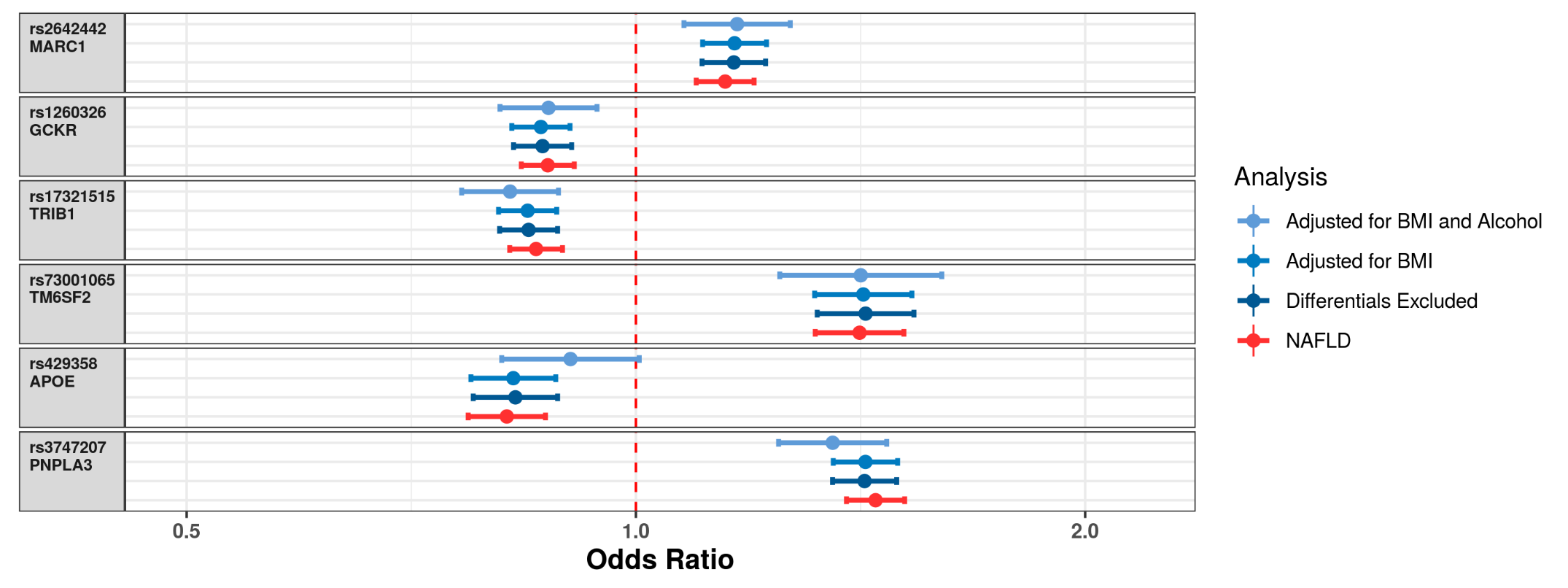

Supplementary Figure 6: Comparison of effect sizes at each lead SNP between the main GWAS and the subgroup analyses. Each point represents the log odds for for the relevant analysis and the error bar represents the 95% confidence interval. The first subgroup analysis excluded any alternative hepatic or alcohol-related pathology. The second subgroup analysis additionally adjusted for body mass index. The third subgroup analysis additionaly adjusted for alcohol intake.

Supplementary Figure 7: Comparison of effect sizes at each lead SNP between the main GWAS, the replication cohort and the two published GWAS summary association statistics. Each point represents the log odds for for the study and the error bar represents the 95% confidence interval
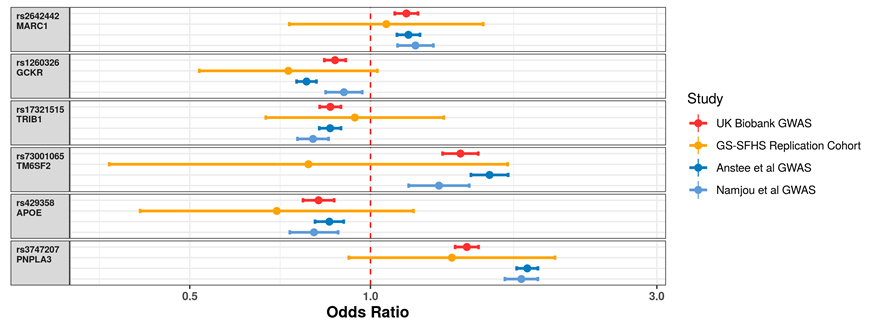
.

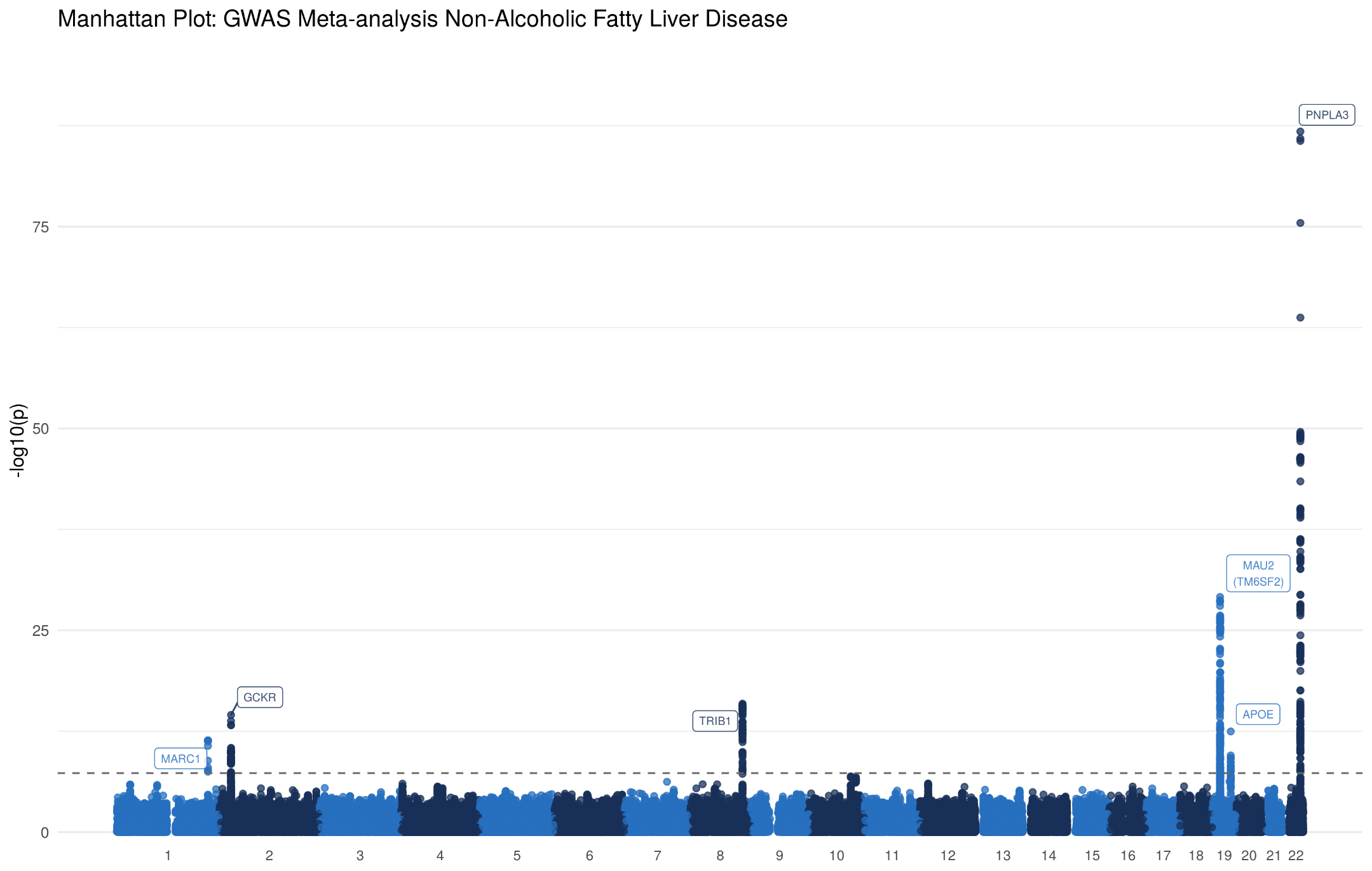

Supplementary Figure 8: Manhattan plot for the association with non-alcoholic fatty liver disease. Meta-analysis of UK Biobank Cohort with data from Namjou et al and Anstee et al. Each variant is plotted based on chromosome and position on the X-axis and -log10 P values on the Y-axis. The horizontal dotted line represents genome-wide significance (5*10^-8^).

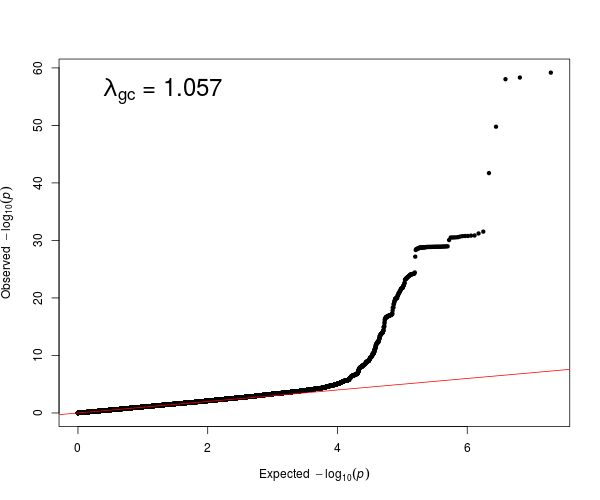

Supplementary Figure 9: QQ Plot showing the observed quantiles of gene effects on Non-Alcoholic Fatty Liver Disease (y-axis) as a function of quantiles expected from a normal distribution (x-axis). Linkage disequilibrium regression score intercept 1.0053.
