## Supplementary Materials 6 for "Genome-Wide Association Study of Non-Alcoholic Fatty Liver Disease Identifies Association with Apolipoprotein E"

### Reporting checklist for genetic association study.

Based on the STREGA guidelines.

|  |  | Reporting Item | Page Number |
| --- | --- | --- | --- |
| **Title and abstract** |  |  |  |
| Title | [#1a](https://www.goodreports.org/reporting-checklists/strega/info/#1a) | Indicate the study’s design with a commonly used term in the title or the abstract | 1 |
| Abstract | [#1b](https://www.goodreports.org/reporting-checklists/strega/info/#1b) | Provide in the abstract an informative and balanced summary of what was done and what was found | 3 |
| **Background/rationale** |  |  |  |
|  | [#2](https://www.goodreports.org/reporting-checklists/strega/info/#2) | Explain the scientific background and rationale for the investigation being reported | 5-6 |
| **Objectives** |  |  |  |
|  | [#3](https://www.goodreports.org/reporting-checklists/strega/info/#3) | State specific objectives, including any prespecified hypotheses. State if the study is the first report of a genetic association, a replication effort, or both. | 5-6 |
| **Study design** |  |  |  |
|  | [#4](https://www.goodreports.org/reporting-checklists/strega/info/#4) | Present key elements of study design early in the paper | 7-14 |
| **Setting** |  |  |  |
|  | [#5](https://www.goodreports.org/reporting-checklists/strega/info/#5) | Describe the setting, locations, and relevant dates, including periods of recruitment, exposure, follow-up, and data collection | 7-10 |
| **Eligibility criteria** |  |  |  |
|  | [#6a](https://www.goodreports.org/reporting-checklists/strega/info/#6a) | Cohort study – Give the eligibility criteria, and the sources and methods of selection of participants. Describe methods of follow-up. Case-control study – Give the eligibility criteria, and the sources and methods of case ascertainment and control selection. Give the rationale for the choice of cases and controls. Cross-sectional study – Give the eligibility criteria, and the sources and methods of selection of participants. Give information on the criteria and methods for selection of subsets of participants from a larger study, when relevant. | 7,8 |
|  | [#6b](https://www.goodreports.org/reporting-checklists/strega/info/#6b) | Cohort study – For matched studies, give matching criteria and number of exposed and unexposed. Case-control study – For matched studies, give matching criteria and the number of controls per case. | 8,9,15 |
| **Variables** |  |  |  |
|  | [#7a](https://www.goodreports.org/reporting-checklists/strega/info/#7a) | Clearly define all outcomes, exposures, predictors, potential confounders, and effect modifiers. Give diagnostic criteria, if applicable | 8-10, Supplement 1 |
|  | [#7b](https://www.goodreports.org/reporting-checklists/strega/info/#7b) | Clearly define genetic exposures (genetic variants) using a widely-used nomenclature system. Identify variables likely to be associated with population stratification (confounding by ethnic origin). | 10,11 |
| **Data sources/measurement** |  |  |  |
|  | [#8a](https://www.goodreports.org/reporting-checklists/strega/info/#8a) | For each variable of interest give sources of data and details of methods of assessment (measurement). Describe comparability of assessment methods if there is more than one group. Give information separately for for exposed and unexposed groups if applicable. | 8-10, Supplement 6 |
|  | [#8b](https://www.goodreports.org/reporting-checklists/strega/info/#8b) | Describe laboratory methods, including source and storage of DNA, genotyping methods and platforms (including the allele calling algorithm used, and its version), error rates and call rates. State the laboratory / centre where genotyping was done. Describe comparability of laboratory methods if there is more than one group. Specify whether genotypes were assigned using all of the data from the study simultaneously or in smaller batches. | 9,10 |
| **Bias** |  |  |  |
|  | [#9a](https://www.goodreports.org/reporting-checklists/strega/info/#9a) | Describe any efforts to address potential sources of bias | 10-14 |
|  | [#9b](https://www.goodreports.org/reporting-checklists/strega/info/#9b) | Describe any efforts to address potential sources of bias | 10-14 |
| **Study size** |  |  |  |
|  | [#10](https://www.goodreports.org/reporting-checklists/strega/info/#10) | Explain how the study size was arrived at | 7,8 |
| **Quantitative variables** |  |  |  |
|  | [#11](https://www.goodreports.org/reporting-checklists/strega/info/#11) | Explain how quantitative variables were handled in the analyses. If applicable, describe which groupings were chosen, and why. If applicable, describe how effects of treatment were dealt with. | 10,12,13 |
| **Statistical methods** |  |  |  |
|  | [#12a](https://www.goodreports.org/reporting-checklists/strega/info/#12a) | Describe all statistical methods, including those used to control for confounding. State software version used and options (or settings) chosen. | 10-14, Supplement 5 |
|  | [#12b](https://www.goodreports.org/reporting-checklists/strega/info/#12b) | Describe any methods used to examine subgroups and interactions | NA |
|  | [#12c](https://www.goodreports.org/reporting-checklists/strega/info/#12c) | Explain how missing data were addressed | NA |
|  | [#12d](https://www.goodreports.org/reporting-checklists/strega/info/#12d) | If applicable, explain how loss to follow-up was addressed | NA |
|  | [#12e](https://www.goodreports.org/reporting-checklists/strega/info/#12e) | Describe any sensitivity analyses | 11-14 |
|  | [#12f](https://www.goodreports.org/reporting-checklists/strega/info/#12f) | State whether Hardy-Weinberg equilibrium was considered and, if so, how. | 6 |
|  | [#12g](https://www.goodreports.org/reporting-checklists/strega/info/#12g) | Describe any methods used for inferring genotypes or haplotypes | 12 |
|  | [#12h](https://www.goodreports.org/reporting-checklists/strega/info/#12h) | Describe any methods used to assess or address population stratification. | 10,12 |
|  | [#12i](https://www.goodreports.org/reporting-checklists/strega/info/#12i) | Describe any methods used to address multiple comparisons or to control risk of false positive findings. | 11 |
|  | [#12j](https://www.goodreports.org/reporting-checklists/strega/info/#12j) | Describe any methods used to address and correct for relatedness among subjects | 10 |
| **Participants** |  |  |  |
|  | [#13a](https://www.goodreports.org/reporting-checklists/strega/info/#13a) | Report numbers of individuals at each stage of study—eg numbers potentially eligible, examined for eligibility, confirmed eligible, included in the study, completing follow-up, and analysed. Give information separately for for exposed and unexposed groups if applicable. Report numbers of individuals in whom genotyping was attempted and numbers of individuals in whom genotyping was successful. | Figure 1 |
|  | [#13b](https://www.goodreports.org/reporting-checklists/strega/info/#13b) | Give reasons for non-participation at each stage | Figure 1 |
|  | [#13c](https://www.goodreports.org/reporting-checklists/strega/info/#13c) | Consider use of a flow diagram | Figure 1 |
| **Descriptive data** |  |  |  |
|  | [#14a](https://www.goodreports.org/reporting-checklists/strega/info/#14a) | Give characteristics of study participants (eg demographic, clinical, social) and information on exposures and potential confounders. Give information separately for exposed and unexposed groups if applicable. Consider giving information by genotype | Supplement 6 |
|  | [#14b](https://www.goodreports.org/reporting-checklists/strega/info/#14b) | Indicate number of participants with missing data for each variable of interest | NA |
|  | [#14c](https://www.goodreports.org/reporting-checklists/strega/info/#14c) | Cohort study – Summarize follow-up time, e.g. average and total amount. | 15 |
| **Outcome data** |  |  |  |
|  | [#15](https://www.goodreports.org/reporting-checklists/strega/info/#15) | Cohort study Report numbers of outcome events or summary measures over time.Give information separately for exposed and unexposed groups if applicable. Report outcomes (phenotypes) for each genotype category over time Case-control study – Report numbers in each exposure category, or summary measures of exposure.Give information separately for cases and controls . Report numbers in each genotype category. Cross-sectional study – Report numbers of outcome events or summary measures. Give information separately for exposed and unexposed groups if applicable. Report outcomes (phenotypes) for each genotype category | Supplement 2 |
| **Main results** |  |  |  |
|  | [#16a](https://www.goodreports.org/reporting-checklists/strega/info/#16a) | Give unadjusted estimates and, if applicable, confounder-adjusted estimates and their precision (eg, 95% confidence interval). Make clear which confounders were adjusted for and why they were included | Tables 1 and 2 |
|  | [#16b](https://www.goodreports.org/reporting-checklists/strega/info/#16b) | Report category boundaries when continuous variables were categorized | NA |
|  | [#16c](https://www.goodreports.org/reporting-checklists/strega/info/#16c) | If relevant, consider translating estimates of relative risk into absolute risk for a meaningful time period | NA |
|  | [#16d](https://www.goodreports.org/reporting-checklists/strega/info/#16d) | Report results of any adjustments for multiple comparisons | Tables 1 and 2, Supplement 2 |
| **Other analyses** |  |  |  |
|  | [#17a](https://www.goodreports.org/reporting-checklists/strega/info/#17a) | Report other analyses done—e.g., analyses of subgroups and interactions, and sensitivity analyses | 17-19, Supplements 2, 4 and 5 |
|  | [#17b](https://www.goodreports.org/reporting-checklists/strega/info/#17b) | Report other analyses done—e.g., analyses of subgroups and interactions, and sensitivity analyses | 17-19, Supplements 2, 4 and 5 |
|  | [#17c](https://www.goodreports.org/reporting-checklists/strega/info/#17c) | Report other analyses done—e.g., analyses of subgroups and interactions, and sensitivity analyses | 17-19, Supplements 2, 4 and 5 |
| **Key results** |  |  |  |
|  | [#18](https://www.goodreports.org/reporting-checklists/strega/info/#18) | Summarise key results with reference to study objectives | 20,21 |
| **Limitations** |  |  |  |
|  | [#19](https://www.goodreports.org/reporting-checklists/strega/info/#19) | Discuss limitations of the study, taking into account sources of potential bias or imprecision. Discuss both direction and magnitude of any potential bias. | 22 |
| **Interpretation** |  |  |  |
|  | [#20](https://www.goodreports.org/reporting-checklists/strega/info/#20) | Give a cautious overall interpretation considering objectives, limitations, multiplicity of analyses, results from similar studies, and other relevant evidence. | 23 |
| **Generalisability** |  |  |  |
|  | [#21](https://www.goodreports.org/reporting-checklists/strega/info/#21) | Discuss the generalisability (external validity) of the study results | 22,23 |
| **Funding** |  |  |  |
|  | [#22](https://www.goodreports.org/reporting-checklists/strega/info/#22) | Give the source of funding and the role of the funders for the present study and, if applicable, for the original study on which the present article is based | 24, Footnotes |

Notes:

- 14a: Supplement 6
- 15: Supplement 2
- 16d: Tables 1 and 2,Supplement 2
- 17a: 10, Supplement 4
- 17b: 10, Supplement 2, Supplement 6
- 17c: 10,11. Supplement 5

The STREGA checklist is distributed under the terms of the Creative Commons Attribution License CC-BY. This checklist was completed on 20. April 2021 using <https://www.goodreports.org/>, a tool made by the [EQUATOR Network](https://www.equator-network.org) in collaboration with [Penelope.ai](https://www.penelope.ai)
