## Supplementary Materials 8 for "Genome-Wide Association Study of Non-Alcoholic Fatty Liver Disease Identifies Association with Apolipoprotein E"

Supplementary Data: Gallstone Diagnostic Codes

Table 1: Diagnostic codes relating to Non-alcoholic fatty liver disease (NAFLD)

Table 2: Diagnostic codes relating to other hepatic pathology, alcohol abuse and diseases relating to alcohol excess

Abbreviations:

ICD9 / ICD10: International Classification of Diseases Ninth and Tenth Revisions

UKB – UK Biobank

READ V2 and READ V3 – Clinical terminology system used in UK Primary Care settings (created by Dr James Read)

OPCS3 and OPCS4 – Office of Population Censuses and Surveys Classification of Interventions and Procedures version 4

| Table 1: NAFLD-Related Pathology: Disease Codes | | | | | |
| --- | --- | --- | --- | --- | --- |
| **Diagnosis** | **ICD10** | **ICD9** | **UKB Code** | **READ V2 Code** | **Additional READ V3 Codes*^1^*** |
| **Non-alcoholic Fatty Liver Disease** | **K75.8, K76.0** | **571.5** | **-** | **C32y5, J6154, J61y1, J61y7, J61y8, J61y9** | **X307v, XaQIT** |
| ***^1^*All READ V2 Codes also apply to READ V3 Codes** | | | | | |

| Table 2: Other Hepatic Pathology: Disease Codes | | | |
| --- | --- | --- | --- |
| **Diagnosis** | **ICD10** | **ICD9** | **UK Biobank Code** |
| **Alcoholic Liver Disease** | **K70** | **571.0-571.3** |  |
| **Viral Hepatitis** | **B16-B19** | **070** | **1156, 1579-1581** |
| **Autoimmune Liver Disease** | **K83.0A, K83.0F, K74.3, K75.4** | **571.6, 576.1** | **1157, 1475** |
| **Haemochromatosis** | **E83.1** | **275.0** | **1507** |
| **Wilson's Disease** | **E83.02** | **275.1** |  |
| **Alpha-1-Antitrypsin Deficiency** | **E88.01, E88.02** | **277.6** | **Direct interview question, 1496** |
| **Budd-Chiari** | **I82.0, K76.5** | **453.0** |  |
| **Chronic Hepatitis Unspecified** | **K73.2, K73.9** | **571.4** | **1155** |
| **Biliary Cirrhosis** | **K74.4, K74.5** | **571.6** | **1159, 1506** |
| **Alcoholic Use Disorder and Complications** | **F10, E24.4, G62.1, I42.6, K29.2, G31.2, G72.1, K85.2, K86.0, T51.0, T51.9, Y57.3, X65, Z50.2, Z71.4, Z72.1** | **303, 305.0, 291, 357.5, 425.5, 535.3, 980.1, 980.9** | **1408, 1604** |
